# A founder population reveals genetic variation with major effect on serious mental illness

**DOI:** 10.64898/2026.09.25.26363706

**Authors:** Greta Gerdes, Veronica Tozzo, Susan K. Service, Clara Frydman-Gani, Ana M. Diaz-Zuluaga, Johanna Valencia-Echeverry, Alejandro Arias, Lucy K. Bicks, Ana M. Ramirez-Diaz, Julia M. Sealock, Calwing Liao, Daniel P. Howrigan, Christiana Liu, Esteban A. Lopera-Maya, Aditya Pimplaskar, Juan F. De la Hoz, Mario A. Arango-Gómez, Camila Arango-Restrepo, Maria C. Arbelaez-Herrera, Katherine Camacho-Gualteros, Mateo Cardona-Arango, Mauricio Castaño-Ramirez, Jaime A. Castañeda-Hoyos, Felecia Cerrato, Sinéad B. Chapman, Roby E. Franco-Gómez, John D. Londoño-Martinez, John S. Mazo-Morales, Juan C. Mejía-Piedrahita, Emaad Mir, Tyler M. Moore, Luz E. Perez-Jaramillo, Maria Pérez-Vallejo, Maria E. Posada-Ricardo, Vladimir Riscanevo-Posada, Camila Restrepo-Osorio, Daniela Restrepo-Osorio, Kosha Ruparel, Danilo Sánchez-Patiño, Terri Teshiba, Jonathan Valdez, Wendy Valencia-Londoño, Oscar A. Valencia-Zuluaga, Jorge M. Velez-Arango, Nicolas A. Crossley, Juan David Palacio-Ortiz, Daniel H. Geschwind, Alicia R. Martin, Ruben C. Gur, Michael R. Sawaya, David S. Eisenberg, Javier I. Escobar, Chiara Sabatti, Victor I. Reus, Carrie E. Bearden, Benjamin M. Neale, Carlos Lopez-Jaramillo, Nelson B. Freimer, Loes M. Olde Loohuis

**Affiliations:** Center for Neurobehavioral Genetics, Semel Institute for Neuroscience and Human Behavior, University of California Los Angeles, Los Angeles, California, USA; Department of Computational Medicine, David Geffen School of Medicine, UCLA, Los Angeles, California, USA; BD2: Breakthrough Discoveries for Thriving with Bipolar Disorder, Santa Monica, California, USA; Research Group in Psychiatry, Department of Psychiatry, School of Medicine, Universidad de Antioquia, Medellín, Antioquia, Colombia; Program in Neurogenetics, Department of Neurology, David Geffen School of Medicine, University of California Los Angeles, Los Angeles, CA, USA; Analytic and Translational Genetics Unit, Department of Medicine, Massachusetts General Hospital, Boston, Massachusetts, USA; Stanley Center for Psychiatric Research, The Broad Institute of MIT and Harvard, Cambridge, Massachusetts, USA; Program in Medical and Population Genetics, The Broad Institute of MIT and Harvard, Cambridge, Massachusetts, USA; Center for Precision Psychiatry, Massachusetts General Hospital, Harvard Medical School, Boston, Massachusetts, USA; Center for Genomic Medicine, Massachusetts General Hospital, Harvard Medical School, Boston, Massachusetts, USA; Department of Mental Health and Human Behavior, Universidad de Caldas, Colombia; Data Engineering Lead, Center for Biosciences, SURA, Colombia; Department of Psychiatry, Perelman School of Medicine, University of Pennsylvania, Philadelphia, PA, USA; Lifespan Brain Institute (LiBI) of Penn Medicine and CHOP, University of Pennsylvania, Philadelphia, PA, USA; Director of Epidemiology and Data Science, Center for Biosciences, SURA, Colombia; Neurodevelopment and Psychosis Section, Department of Psychiatry, University of Pennsylvania, Philadelphia, PA, USA; Center for Biosciences, SURA, Colombia; Department of Psychiatry, Pontificia Universidad Católica de Chile, Santiago, Chile; Centro de Interés Nacional para Investigación e Innovación en Niñez, Adolescencia, Resiliencia y Adversidad (IINARA), Santiago, Chile; Department of Psychiatry, School of Medicine, Universidad de Antioquia, Medellín, Antioquia, Colombia; Center for Autism Research and Treatment, Semel Institute, David Geffen School of Medicine, University of California Los Angeles, Los Angeles, CA, USA; Department of Human Genetics, David Geffen School of Medicine, University of California Los Angeles, Los Angeles, CA, USA; Institute for Precision Health, University of California Los Angeles, Los Angeles, CA, USA; Brain Behavior Laboratory, Department of Psychiatry, Perelman School of Medicine, University of Pennsylvania, Philadelphia, PA, USA; 25 Department of Biological Chemistry, University of California, Los Angeles, CA, USA; 26 Department of Chemistry and Biochemistry, University of California, Los Angeles, CA, USA; Department of Energy Institute for Genomics and Proteomics, University of California, Los Angeles, CA, USA; 28 Molecular Biology Institute, University of California, Los Angeles, CA, USA; Rutgers University, New Brunswick, NJ, USA; Department of Biomedical Data Science, Stanford, CA, USA; Department of Psychiatry and Behavioral Sciences, School of Medicine, University of California, San Francisco, San Francisco, California, USA; Department of Psychology, University of California Los Angeles, Los Angeles, California, USA; Department of Psychiatry, David Geffen School of Medicine, University of California Los Angeles, Los Angeles, California, USA

## Abstract

Genetic studies of serious mental illness (SMI, including schizophrenia and bipolar disorder) have implicated two classes of risk variant: common variants^1–3^, each of small effect but collectively accounting for most population-level risk, and ultra-rare coding variants^4,5^ which confer large risk in a small number of individuals. Most such studies cannot directly assess the impact of specific variants on SMI, transdiagnostically, as the phenotypic information that they collect is focused on a single diagnosis. Here we report results of association analysis of predicted deleterious variants from exome sequencing of 21,958 cases (SMI and severe major depressive disorder) and 19,826 controls from the Mision Origen biobank in the Paisa genetic isolate of Colombia. We identified transdiagnostic associations to seven genes, three at an exome-wide and four at a false discovery rate significance threshold. Five of these genes have not previously been implicated in SMI (*ADAP1*, *WIPI1*, *NFE2L1*, *FBXO10*, and *HLA-DPB1*); for the first four of these the association is driven by variants enriched in the Paisa due to a founder effect, filling the gap between ultra-rare and common variants. The strongest association that we observe is to a frameshift variant (7:904169:T:TC) in *ADAP1* that is genome-wide significant for SMI, schizophrenia, and bipolar disorder and increases risk for both diagnoses by more than threefold; it truncates ADAP1, a brain-enriched Arf6 GTPase-activating/scaffolding protein that has been mechanistically linked to post-mitotic neuronal morphogenesis and regulation of dendritic differentiation^6^. Variant carriers differ significantly from other SMI cases in Mision Origen for psychotic, manic, and cognitive item-level phenotypes assessed through analyses of electronic health records^7–9^ and displayed impaired executive function on neurocognitive testing. Variant carriers differ significantly from other SMI cases in Mision Origen for multiple item-level phenotypes (psychotic, manic, and cognitive) assigned through natural language processing of longitudinal electronic health records. Although the 7:904169:T:TC variant is not significantly associated with major depressive disorder, carriers compared to others with this diagnosis display a significant increase in symptoms typical of bipolar disorder. The variant’s high allele frequency in Mision Origen (∼0.4%) will enable population-level studies to stratify SMI based on causation and identify additional factors influencing risk and resilience.

## Main

Mental health research and clinical practice classify serious mental illness (SMI) according to discrete diagnoses, characterized by either the predominance of psychotic symptoms (schizophrenia and schizoaffective disorder [SCZ]) or disturbances of mood (the rubric “mood disorders includes bipolar disorders [BD]^10,11^ but also frequently incorporates unipolar major depressive disorder [MDD]). This classification system, introduced in the early 20th Century^12^, remains the international standard despite strong and growing evidence that these categories are both over-specific (symptoms overlap considerably between diagnoses) and under-specific (each diagnosis encompasses substantial phenotypic heterogeneity^13,14^). Genetic studies of both rare and common variants have adopted this framework, focusing on identifying loci and genes associated with individual diagnoses rather than SMI as a unified spectrum. Cross-disorder analyses, however, have shown that most of the genetic variance identified for individual diagnoses is best represented by transdiagnostic factors^15^. Such analyses have required aggregation of data from multiple different studies, encompassing diverse approaches for assessing and designating diagnoses; as few of these studies have recruited cases across SMI categories or systematically recorded information on sub-diagnostic or dimensional features, transdiagnostic genetic discovery efforts for SMI have been underpowered^16,17^. We created the Mision Origen biobank in Northwestern Colombia as a resource for transdiagnostic discovery of genetic variants with a large impact on risk for SMI, enabled by two distinctive advantages of this region and its predominant population (referred to as “the Paisa”).

First, most hospitals in the Paisa region maintain extensive longitudinal electronic health record (EHR) databases. We have used the EHR of four specialty psychiatric hospitals to uniformly ascertain, recruit, and phenotype individuals with EHR diagnoses of SCZ (including schizoaffective disorder, which we group with SCZ for all analyses reported here), BD, and severe MDD (**Table S1**). We have recruited controls through five primary care clinics operated by SURA, the largest health insurance and health care provider in the region, screening them to exclude the above diagnoses and matching them to cases by age, sex, educational attainment and geographic region of birth. Through systematic analyses of structured EHR data we assigned diagnoses to participants and obtained information on demographics, medication use, and health care utilization. By developing and validating Natural Language Processing (NLP) pipelines, we extracted data to assign sub-diagnostic phenotypes (psychiatric symptoms and behaviors) to all participants using the free text of clinical notes^7–9,18^.

Second, the Paisa population has a history and demography that facilitates discovery of individual damaging genetic variants with a large impact on disease phenotypes. In most populations such single variants are difficult to detect through exome-wide or genome-wide association studies, as they are generally strongly selected against and therefore rare^19^. However, in a population isolate such as the Paisa that has expanded rapidly from a recent bottleneck, such variants may rise to a frequency that is orders of magnitude greater than in other populations if they were present in the population during the early stages of its expansion; this phenomenon has contributed to several discoveries of large-effect associations to common diseases or quantitative traits in different populations^20–25^. The Paisa population is concentrated in valleys situated between the Central and Western ranges of the Andes Mountains and was founded in the 16th and 17th Centuries through admixture between European colonists (mainly male), indigenous Amerindians (mostly female)^26,27^, and enslaved Africans. Over the subsequent four centuries this population grew by more than 100-fold from the founding bottleneck^28^, with minimal further immigration until the beginning of the 20th Century; now, with about 9 million mostly urban inhabitants, the Paisa is one of the largest and most geographically concentrated isolates globally, a fact that facilitated the creation of the Mision Origen biobank. Based on the size of Mision Origen, and indications from prior studies that the Paisa has population genetic characteristics comparable to the other genetic isolates noted above^29–32^, we hypothesized that an exome wide association study of this cohort would be well-powered for discovery of novel significant large-effect associations to SMI disorders across a wide spectrum of variant frequencies.

We performed Blended Genome Exome (BGE) sequencing in 52,848 participants. In this approach a single DNA library is used to generate deep whole exome coverage (average 30-40x) and low-pass whole genome coverage (average 1-4x) in a single sequencing run^33^. After extensive quality filtering of genotypes and phenotypes and the exclusion of related individuals (up to second degree), 41,784 participants were retained for the exome-wide association analyses reported here (n=21,958 cases [SCZ, 3688, BD, 8507; MDD, 9763] and 19,826 controls; **Methods**, **Fig. S1-S3, Table S2**). The mean age of analyzed participants was 48 years (range 18-90) and 63% (n=26,513) were female.

We conducted single-variant analyses of the BGE exome data to discover associations to low-frequency variants (0.1%-1% minor allele frequency [MAF]) and gene-based burden analyses to identify associations to rare (0.01%-0.1% MAF) and ultra rare variants (< 0.01% MAF). We identified large-effect transdiagnostic associations at seven genes (significant at genome-wide/exome-wide [n=3], or False Discovery Rate [FDR] thresholds [n=4]), four of which (*ADAP1*, *WIPI1*, *NFE2L1*, and *FBXO10*) were driven by variants with greatly increased frequency in the Paisa population and have not previously been implicated in any mental disorder. Notably, a protein-truncating insertion variant (7:904169:T:TC) in *ADAP1* (ADP-ribosylation factor GTPase-activating protein [ArfGAP] with dual pleckstrin homology [PH] domains) displays genome-wide significant association with SMI, transdiagnostically, displaying a greater than threefold increased risk for both SCZ and BD. Multiple sub-diagnostic phenotypes are overrepresented among SMI or MDD carriers of this variant, which is greatly enriched in Mision Origen; this observation suggests an unprecedented opportunity to stratify SMI, broadly defined, based on biological causation.

### Identifying large-effect SMI associations

We conducted single-variant association analyses using 21,506 exonic variants (all protein truncating and missense predicted deleterious variants with minor allele count [MAC] ≥3, **Fig. S4**) for five phenotypes: two transdiagnostic (SMI [SCZ and BD] and SMI+ [SMI and MDD]); and three diagnosis specific (SCZ, BD, and MDD, **Table 1a, Fig. S5-S6, Tables S3-S7)**. Our strongest associations were to a protein truncating frameshift insertion variant (7:904169:T:TC) in *ADAP1* with a large effect on SMI, transdiagnostically. This variant is carried by 258 case participants (256 heterozygous, 2 homozygous) and 85 heterozygous controls (**Table S8**) with a global minor allele frequency (MAF) of 0.4% (0.6% in cases vs 0.2% in controls); in the gnomAD database the variant was observed in only two admixed American (AMR) individuals and was not found in individuals from other ancestries.

**Table 1:** Single variant and gene-based associations to serious mental illness in Mision Origen.

(a)
| Variant | Gene | Outcome | Consequence | Significance | OR (95% CI) | P-value | AC cases/<br>controls | MAF<br>Mision<br>Origen | MAF<br>gnomAD<br>AMR | MAF<br>gnomAD<br>EUR |
| --- | --- | --- | --- | --- | --- | --- | --- | --- | --- | --- |
| 7:904169:T:TC | *ADAP1* | SMI | PTV | Genome-wide | 3.33 (2.60, 4.26) | 4.87e-22 | 199/85 | 4.52e-03 | 3.33e-05 | 0 |
|  |  | SCZ |  | Genome-wide | 3.50 (2.37, 5.18) | 1.61e-10 | 47/85 | 2.85e-03 |  |  |
|  |  | BD |  | Genome-wide | 3.26 (2.50, 4.25) | 1.36e-18 | 152/85 | 4.26e-03 |  |  |
|  |  | SMI+ |  | Genome-wide | 2.47 (1.94, 3.15) | 9.16e-14 | 260/85 | 4.21e-03 |  |  |
| 6:33086236:TA:T | *HLA-DPB1* | SMI | PTV | FDR | 1.24 (1.14, 1.35) | 4.75e-07 | 1007/1302 | 3.79e-02 | 3.74e-02 | 2.57e-02 |
| 17:68433486:A:G | *WIPI1* | SMI | Damaging<br>missense | FDR | 4.14 (2.28, 7.53) | 1.62e-06 | 34/12 | 7.32e-04 | 1.67e-05 | 8.48e-07 |

(b)
| Gene | Outcome | Variant<br>inclusion | Significance | OR (95% CI) | P-value | AC<br>cases/controls | Variants MAC>5<br>/Variants MAC≤5 |
| --- | --- | --- | --- | --- | --- | --- | --- |
| *ADAP1* (chr 7) | SMI | All | Exome-wide | 3.00 (2.37, 3.80) | 5.84e-20 | 203/97 | 1/12 |
|  | SCZ | All | Exome-wide | 3.16 (2.17, 4.60) | 1.05e-09 | 49/97 | 1/11 |
|  | BD | All | Exome-wide | 2.94 (2.27, 3.80) | 1.03e-16 | 154/97 | 1/11 |
|  | SMI+ | All | Exome-wide | 2.25 (1.79, 2.83) | 2.93e-12 | 268/97 | 1/15 |
| *WIPI1* (chr 17) | SMI | All | Exome-wide | 4.26 (2.35, 7.17) | 8.67e-07 | 35/12 | 1/1 |
|  | BD | All | FDR | 4.23 (2.23, 8.04) | 5.28e-06 | 26/12 | 1/1 |
| *KDM5B* (chr 1) | SMI | MAC≤5 | Exome-wide | 3.95 (2.29, 6.82) | 4.08e-07 | 39/16 | 0/44 |
|  | BD | MAC≤5 | Exome-wide | 4.82 (2.72, 8.55) | 3.78e-08 | 33/16 | 0/39 |
| *NFE2L1* (chr 17) | SMI | All | FDR | 1.75 (1.35, 2.28) | 1.38e-05 | 123/115 | 1/10 |
| *ZMYM2* (chr 13) | SMI | MAC≤5 | FDR | 39.84 (8.16, 194.65) | 2.64e-06 | 12/0 | 0/12 |
| *FBXO10* (chr 9) | SMI+ | All | FDR | 0.40 (0.27, 0.59) | 2.84e-06 | 32/76 | 3/4 |

a) Single variant associations: Significant associations are listed between predicted deleterious variants (damaging missense and PTV) and both transdiagnostic (SMI and SMI+) and disease specific outcomes (BD, SCZ); MAF are reported for Mision Origen and for the gnomAD v4.1.1 AMR and EUR reference populations<sup>89</sup>. (b) Gene-based (burden) associations: Significant associations are listed between predicted deleterious variants and both transdiagnostic and disease-specific phenotypes, as above. Analyses were performed using two variant sets: all predicted deleterious variants regardless of minor allele count, and a restricted set of ultra-rare variants (MAC ≤ 5). Gene-based association results for *ZMYM2* with SMI (no MAC filter) were excluded from the table as all variants had MAC ≤ 5.
AC, allele count; AMR, admixed American; BD, bipolar disorder; EUR, European; FDR, false discovery rate; MAC, minor allele count; MAF, minor allele frequency; OR, odds ratio; PTV, protein truncating variant; SCZ, schizophrenia; SMI, serious mental illness; SMI+, serious mental illness + major depressive disorder

The 7:904169:T:TC variant is located in exon 6 of *ADAP1* and introduces a downstream stop codon that prematurely truncates ADAP1. Genome-wide significant associations were observed to SMI (odds ratio OR=3.33, 95% CI=[2.60-4.26]; p=4.87e-22), SMI+ (OR=2.47, 95% CI=[1.94-3.15]; p=9.16e-14), SCZ (OR=3.50, 95% CI=[2.37–5.18]; p=1.61e-10) and BD (OR=3.26, 95% CI=[2.50, 4.25]; p=1.36e-18). For MDD the effect size was smaller and the association was non-significant (OR=1.38, 95% CI=[0.98–1.94]; p=6.88e-02), although the direction of effect was consistent with that of the other phenotypes.

The single-variant analyses also identified variants in two genes at which associations to SMI were significant at a False Discovery Rate (FDR) of <5%. One variant, in *HLA-DPB1* is a predicted protein truncating variant (6:33086236:TA:T) that is relatively common (MAF > 1%) in Mision Origen and all major reference populations in gnomAD. The second variant, in *WIPI1* is a predicted damaging missense variant (17:68433486:A:G) present in 46 case participants (45 heterozygous, 1 homozygous) and 12 heterozygous controls in Mision Origen (**Table S8**), and present in gnomAD only once among AMR samples and once among EUR samples; this variant exerts comparable effects on both SCZ and BD risk (**Table 1a, Fig. S5**).

Analyses conditioned on common variant association signals are important for determining that the loci identified through single-variant analyses are independent of such signals. For 7:904169:T:TC, we performed conditional analyses adjusting for lead SNPs, identified in Mision Origen, within a nearby region (7:1,817,033–2,283,308) previously highlighted by genome wide association studies (GWAS) of BD, SCZ and MDD and a cross-diagnosis analysis^1,34^ (see **Methods** and Lopera-Maya et al.^29^). The association of 7:904169:T:TC with SMI-related phenotypes remained strong and significant after conditioning on all nearby common variant signals (**Supplementary results, Fig. S7–S9, Tables S9–S10**). Such analyses were unnecessary for the 17:68433486:A:G variant in *WIPI1*, as there were no common variant results in the surrounding region that were significant in Mision Origen. The single base-pair deletion 6:33086236:TA:T in *HLA-DPB1*, lies in the major histocompatibility complex, which has previously demonstrated associations to multiple mental disorders^1,15,35^. Analysis to place this SMI locus in the context of these previous findings requires a more precise imputation for this complicated genomic region than is currently available.

We performed gene-based burden analyses in the Mision Origen cases and controls, using the same set of transdiagnostic and single-diagnosis phenotypes described above in single-variant tests. For these analyses damaging variants within each gene were aggregated and tested for association with SAIGE-GENE using Firth logistic regression. To enable evaluation of both low frequency/rare and ultra-rare variant effects, we conducted two analyses: one considered all damaging variants regardless of MAC, and one considered only variants with MAC ≤5 (**Tables S11-S20, Fig. S6**). Three genes, *ADAP1*, *WIPI1*, and *KDM5B,* were associated with SMI at an exome-wide significance threshold (**Table 1b, Fig. S10-S11**). For *ADAP1* (OR=3.00, 95% CI=[2.37-3.80]; p=5.84e-20) and *WIPI1* (OR=4.26, 95% CI=[2.35-7.71]; p=8.67e-07) these associations were detected in the analyses that included all deleterious variants regardless of MAC and were driven by the same intermediate-frequency variant highlighted in the single variant analyses. For these genes we also observed associations with other phenotypes: SMI+, SCZ, and BD, for *ADAP1* (all at an exome-wide significance level) and BD for *WIPI1* (at an FDR of < 5%). *KDM5B,* by contrast, reached exome-wide significance in the model including only variants with MAC≤5, both for SMI (OR=3.95, 95% CI=[2.29-6.82]; p=4.08e-07) and BD (**Table 1b, Fig. S10**).

We further identified transdiagnostic gene-based associations to an additional three genes at FDR < 5%; *ZMYM2* and *NFE2L1* were associated with SMI, while *FBXO10* was negatively associated with SMI+; the suggestion of a protective effect for FBXO10 variation should be interpreted cautiously, pending completion of sequencing of a larger sample of MO participants and further characterization of both the cases and controls carrying these variants. For all three genes the associations were observed under a model that included all variants regardless of MAC. Out of the seven genes identified, only *ZMYM2* and *NFE2L1* are intolerant to loss-of-function variation (pLI=1^36^, LOEUF<0.6 for both genes in gnomADv.4.1.1^37^), while the other genes are relatively tolerant to loss-of-function variation (pLI=0, LOEUF>0.6).

### Bridging Rare and Common Variant Findings

The genetic architecture of common diseases can be represented in terms of the reciprocal relationship between the effect size and population frequency of associated variants. Previous knowledge of the specific genes and loci associated with SMI disorders was limited to two distinct regions of this distribution (**Fig. 1a**); large-effect associations to rare and ultra-rare variation -including both burden-test associations to coding variants in highly constrained genes (as shown here) and associations to copy number variants that span dozens to hundreds of genes^38^ - and small-effect GWAS associations to common variants.

**Fig. 1:**
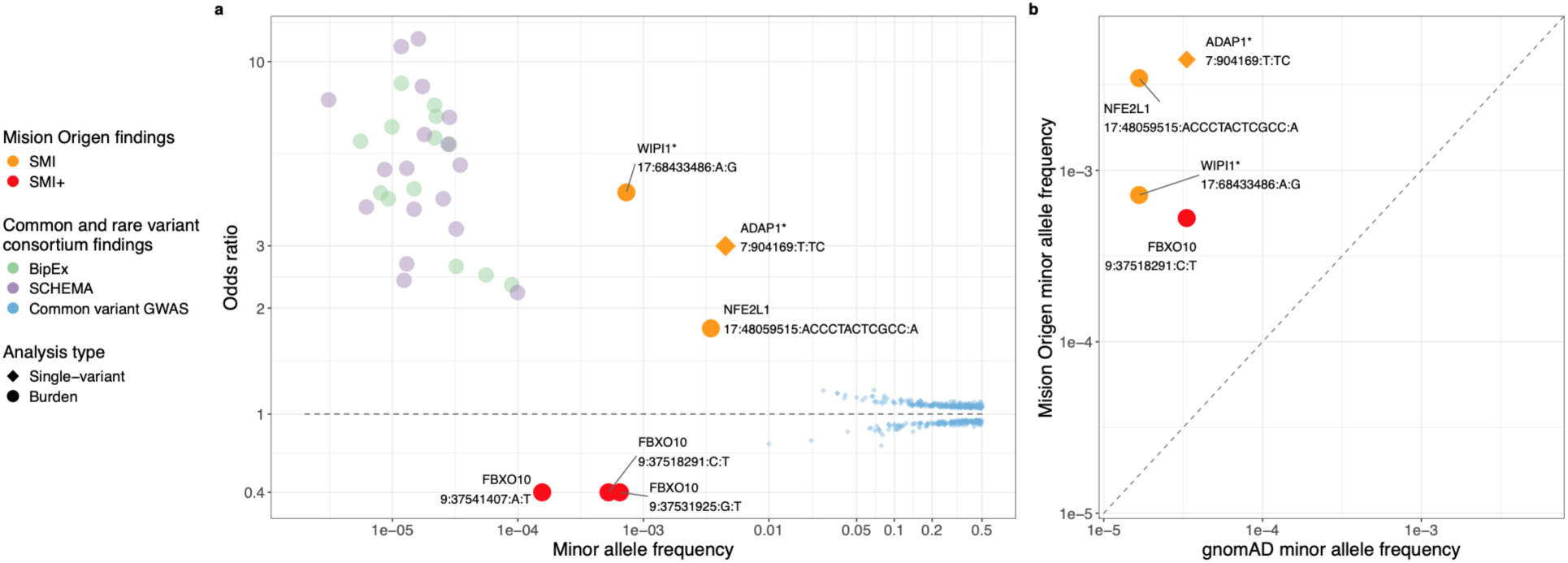
Deleterious variants with increased frequency in the Paisa population are responsible for large-effect transdiagnostic associations. (a) Comparison of Mision Origen associations for SMI or SMI+ with international consortia findings for individual diagnoses in terms of effect size and MAF. Rare variant findings from BipEx2^4^ for BD (green circles) and SCHEMA2^5^ for SCZ (purple circles) depict all genes for which gene-based tests achieved exome-wide significance (for PTV alone or PTV and damaging missense variants combined), including the ORs for those tests and reference MAF values from gnomAD v4.1.1. Common variant findings (genome-wide significant associations for SCZ and BD) are shown as blue dots^1–3^. For Mision Origen, the orange (SMI) or red (SMI+) symbols represent genes for which single-variant (diamonds) or gene-based (circles) tests identified genome-wide/exome wide significant (designated with an asterisk) or FDR significant associations to PTV or damaging missense variants. The variant labels indicate individually significant variants or the lead variants that drove the association signal in gene-based burden analyses (plotted with burden OR), with MAF corresponding to that of the listed variant. (b) Comparison of MAF for individually significant variants (diamonds) or lead variants (Table S21) in gene-based burden analyses (circles) observed in Mision Origen (y-axis) versus gnomAD v4.1.1 (x-axis). The dashed line indicates the identity line (x = y); points above the line indicate higher frequency in Mision Origen relative to gnomAD.

The association findings that we report here (at genome-wide, exome-wide, or FDR significance thresholds) occupy the gap between these two poles. Our results implicating four new genes in the causation of SMI or SMI+ were driven by associations to individual deleterious variants with sizable effects on these phenotypes (**Fig. 1a**): ADAP1 (7:904169:T:TC, SMI OR=3.33); WIPI1 (17:68433486:A:G, SMI OR=4.14); NFE2L1 (17:48059515:ACCCTACTCGCC:A, SMI OR=1.78); FBXO10 (9:37518291:C:T, SMI+ OR=0.45; 9:37531925:G:T, SMI+ OR=0.42; 9:37541407:A:T, SMI+ OR=0.21), see **Table S21**. For each of these variants, the population frequencies (as measured in the Paisa), ranging from 1.56e-04 at 9:37541407:A:T in FBXO10 to 4.43e-03 at 7:904169:T:TC in ADAP1, were intermediate between those of the common variants comprising the GWAS results and the ultra-rare variants comprising recent SCZ (SCHEMA2) and BD (BipEx2) consortium findings^4,5,39^(**Fig. 1a**). The specific allele frequencies and effect sizes are likely to be influenced by winner’s curse as may become evident in further sequencing studies in this population.

For the other two genes at which we found significant associations to SMI in Mision Origen (*KDM5B* and *ZMYM2*), the burden test results reflect the effects of ultra-rare variants, as also identified for these genes in BipEx2^4^ and SCHEMA2^5^, and large-scale autism sequencing studies^40^. Of the remaining genes that reached exome-wide significance in the consortium studies (as shown in **Fig. 1a**) none achieved significance in our dataset, which is less than 1/5 the size of the consortium datasets. Most of these genes however, displayed associations in our dataset with effect sizes roughly comparable to those in SCHEMA and BipEx (**Fig. S12-S13**), consistent with prior evidence for the conservation of ultra-rare variant signals for SMI disorders across populations^39^; one caveat to this interpretation is that Mision Origen case and control samples were included in the most recent consortium analyses (n=19,424 participants in SCHEMA2, 8% of the total sample; and n=27,378 participants in BipEx2,12% of the total sample).

The most parsimonious explanation for our discovery of associations at genes not previously implicated in SMI disorders is that, as in other recently expanded genetic isolates, random genetic drift during the rapid expansion of the Paisa population enabled variants that passed through its founding bottleneck to attain a dramatically increased frequency in this population compared to others. This explanation is consistent with the sparsity or absence of these variants in gnomAD or related databases (see **Fig. 1b, Table S22**). It is important to note, however, that the populations that contributed to the founding Paisa population (Iberian, Indigenous American from Colombia, and African) are themselves relatively sparsely represented in gnomAD and other public sequence databases. Additionally, the history of tri-continental admixture prior to its expansion distinguishes the Paisa from other large isolates, and raises the possibility that population genetic processes other than random drift (e.g., balancing selection of variants introduced to South America by immigrants) could have led to increased frequency of variants associated with SMI; this possibility motivates efforts to explore the ancestry of such variants.

### Paisa history shapes genetic findings

We focused here on identifying the likely continental source of 7:904169:T:TC and 17:68433486:A:G, the two variants overrepresented in Mision Origen that displayed the strongest individual associations with SMI. Using RFMIX^41^, we computed 3-way local ancestry across the phased first 20megabases of chromosome 7 (positions 0–20,000,000) and the 20-megabase region surrounding 17:68433486:A:G (positions 58,433,486-78,433,486). Evaluation of the most likely haplotypes containing the respective variants (from positions 34,713–924,756 on chromosome 7 and 68,379,734–68,557,574 on chromosome 17) indicates that both variants likely originated on a chromosome with European (EUR) ancestry; such ancestry is predicted for 344/345 carrier haplotypes of 7:904169:T:TC with the remaining carrier haplotype predicted as having AMR ancestry; and all 59/59 carrier haplotypes of 17:68433486:A:G carriers. It is not yet evident whether these variants originated in Europe or arose on a European background after the Spanish conquest of what is now Colombia in the 16th Century. Their absence in European sequence databases (**Table S22**) suggests the latter explanation, however as noted above, such databases currently have sparse representation from Iberian populations. Local ancestry-informed association analysis using Tractor-Mix^42^ confirmed that the association signals at both loci are driven exclusively by EUR ancestry haplotypes, with confidence intervals of effect sizes overlapping those obtained through ancestry-agnostic analysis (**Supplementary Results, Tables S23-S24**).

The frequency of the 7:904169:T:TC *ADAP1* variant and the 17:68433486:A:G *WIPI1* variant among Mision Origen control participants suggests that tens of thousands of individuals in the Paisa region may be carrying these variants, most of whom are unlikely to carry an SMI diagnosis. Evaluating population-level risk at this site is an important focus for future research, and conceivably for future clinical practice. As a first step in investigating the generalizability of these findings within the Paisa population, we examined 7:904169:T:TC and 17:68433486:A:G in terms of their geographical clustering and the relative genetic relatedness of their carriers. To quantify spatial clustering, we computed Moran’s I, a measure of spatial autocorrelation that ranges from -1 (perfectly dispersed) to +1 (perfectly clustered), with values near 0 indicating random geographic distribution. Carriers of both variants showed modest but significant geographic clustering relative to non-carriers at the municipality level (7:904169:T:TC: Moran’s I=0.08, p=1.75e-02; 17:68433486:A:G I=0.06, p=3.69e-02), based on 1,000,000 Monte Carlo simulations (**Supplementary Results**). **Fig. 2** summarizes Mision Origen recruitment and geographic distribution of 7:904169:T:TC variant carriers (see **Fig. S14** for 17:68433486:A:G). Consistent with a shared founder for each variant, variant carriers were also significantly more genetically related to one another than non-carrier pairs, despite removal of closely related individuals from all analyses: 10.8% of 7:904169:T:TC carrier pairs and 3.0% of 17:68433486:A:G carrier pairs were related at levels between second and first cousin, compared to only 1% of non-carrier pairs (p<1e-06 and p=2.58e-02 respectively, based on 1 million permutations using PLINK’s Identity-by-State test procedure, see **Supplementary Results**).

**Fig. 2:**
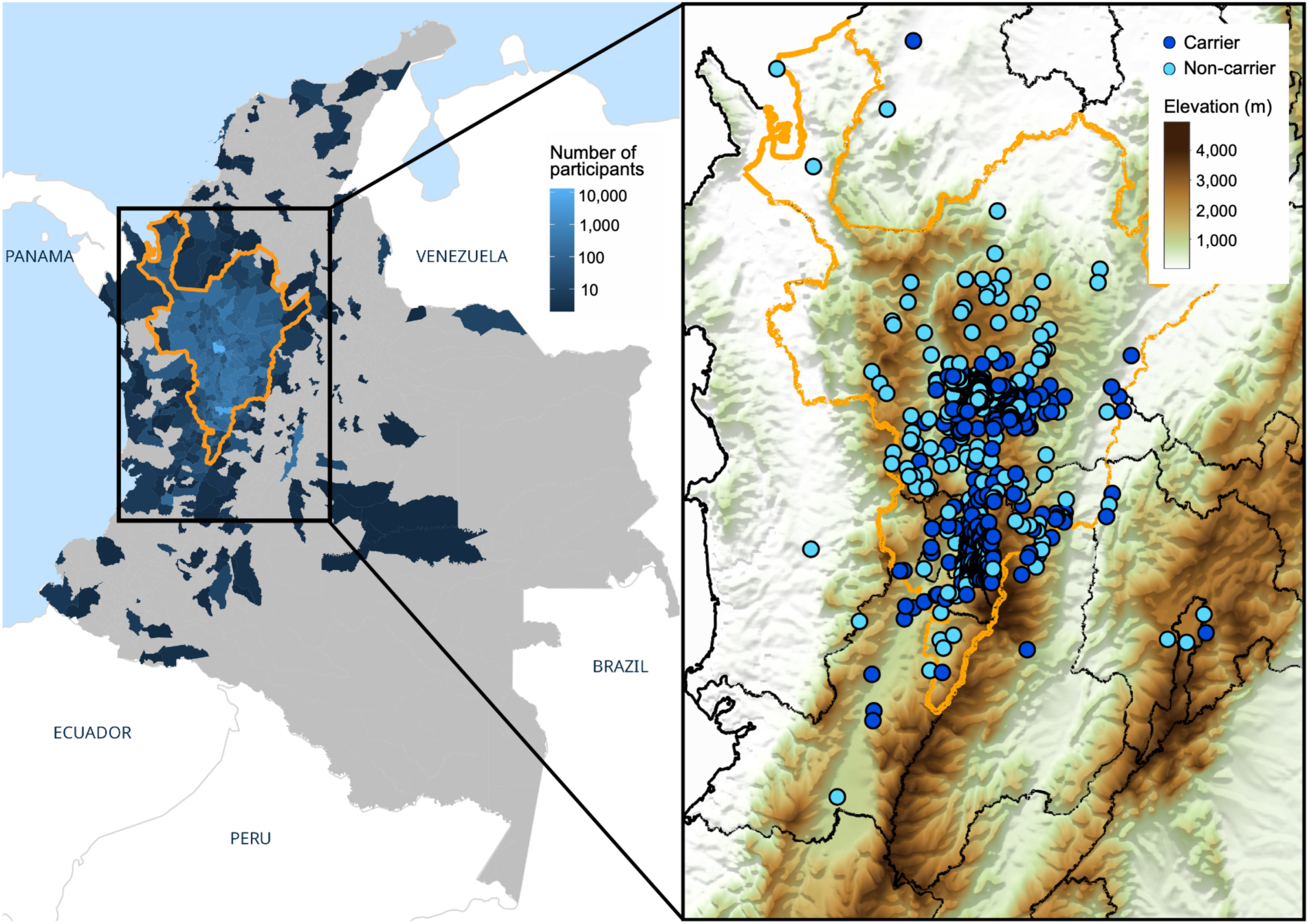
The 7:904169:T:TC variant shows geographical clustering in the Paisa region of Colombia. Geographic distribution of study participants’ place of birth (left) shown as number of participants included in genetic analyses per municipality. Color intensity reflects the number of participants per municipality; municipalities with no registered participants are shown in gray. The Paisa region is outlined in orange. Geographic distribution of 7:904169:T:TC variant carriers and equal number of randomly selected non-carriers (right). To protect participant confidentiality, locations are jittered around city centers and do not represent individual residential addresses or exact birth locations. Positions are overlaid on a topographic relief map of the Andean mountain range.

### ADAP1 and WIPI1 gene expression

*ADAP1* and *WIPI1* are the two genes with significant associations in Mision Origen whose tissue-specific or developmental expression patterns have not been described in SCHEMA2 or BIPEX2. To place ADAP1 and WIPI1 in cellular and developmental context, we examined their expression across cell types and development. In adult human cortex (Wamsley et al., 2024^43^) and whole-brain (Siletti et al., 2023^44^) both ADAP1 and WIPI1 were broadly expressed across cell types and brain regions at low levels **(Fig 3a**). Across development, both ADAP1 and WIPI1 showed increasing expression following birth, with ADAP1 expression peaks around 1 year after birth and WIPI1 expression continues to increase across childhood, peaking around age 10, and both genes are highly expressed in adults, during the peak periods of onset of SMI (**Fig 3b**). Protein-protein interacting partners showed enrichment in biological properties including peptidyl serine modification and cytoskeleton organization for ADAP1 (**Fig 3c-d)** and vacuole organization, autophagosome organization, and mitochondrion organization for WIPI1 (**Fig 3c-d**).

**Fig. 3:**
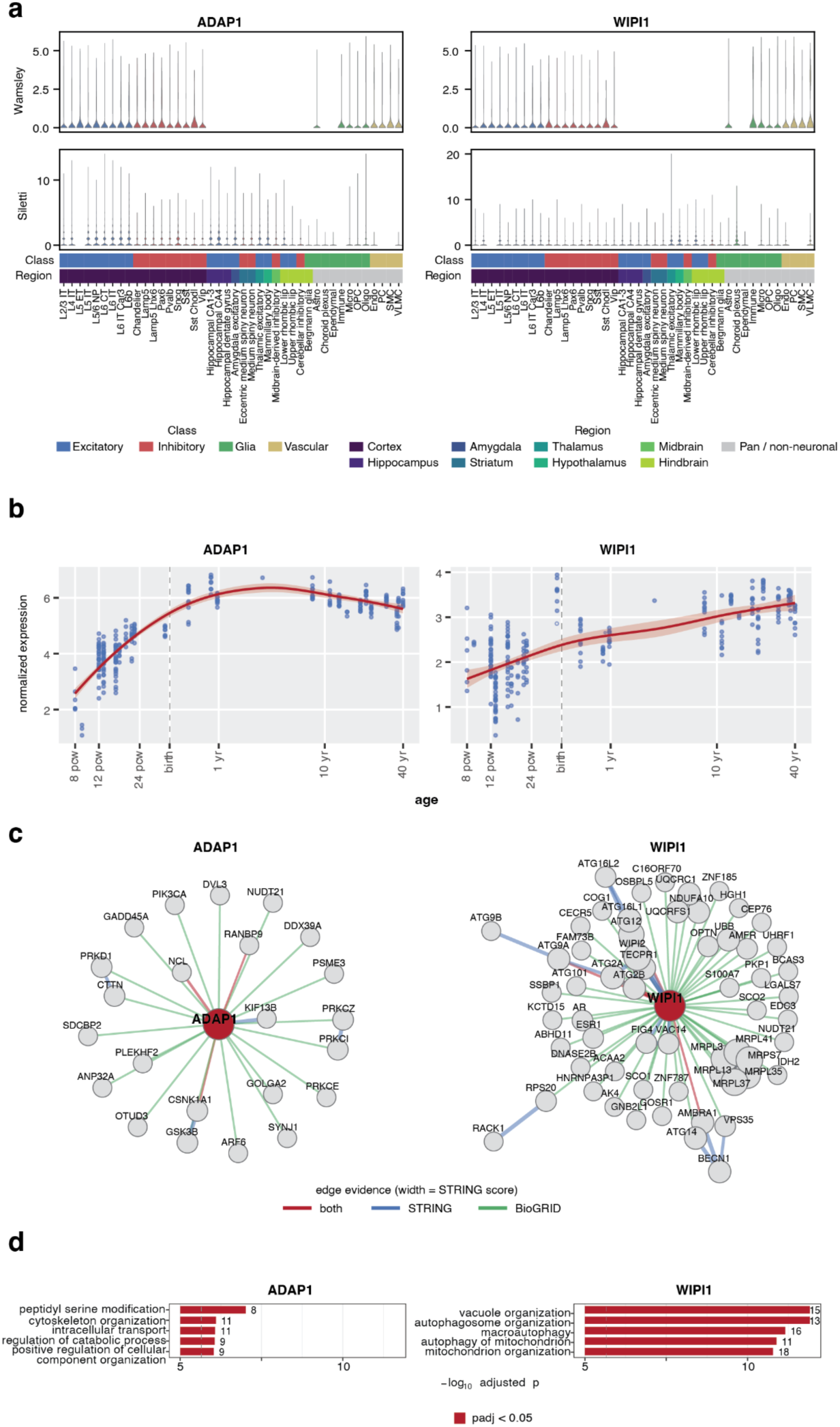
Gene expression and protein interaction patterns of ADAP1 and WIPI1. Expression of ADAP1 (left) and WIPI1 (right) across cell types in adult human cortex (Wamsley et al.^43^, 2024; top) and whole brain (Siletti et al., 2023^44^; bottom). Cell types are ordered by region and major class, indicated by annotation bars (Class: excitatory, inhibitory, glia, vascular; Region: cortex, hippocampus, amygdala, striatum, thalamus, hypothalamus, midbrain, hindbrain, and pan/non-neuronal). b) Developmental trajectories in BrainSpan bulk RNA-seq of human cortex. Points are individual samples; dashed vertical line represents birth. c) Protein-protein interaction networks for ADAP1 (left) and WIPI1 (right), taken as the union of BioGRID and STRING physical interactions. Edge color indicates supporting evidence; width indicates STRING confidence score (min score = 0.7). d) Top five over-represented GO biological process terms among each interactor set. Dashed line marks adjusted p = 0.05.

### The 7:904169:T:TC variant stratifies SMI and MDD

Because detailed longitudinal EHR data were available for most Mision Origen participants, we tested whether comparing item-level clinical phenotypes between carriers and non-carriers could reveal clinically meaningful stratification within SMI diagnoses. We focused these analyses on the *ADAP1* 7:904169:T:TC variant given the substantially larger sample of SMI carriers (n=198) compared to other variants overrepresented in the Paisa region, the strength of its genome-wide association, and the substantial number of MDD carriers (n= 60) enabling us to also evaluate if it stratifies this phenotype.

We analyzed all symptoms extracted from psychiatrist notes using Natural Language Processing software and structured EHR-derived features using EHR data available from three of the four case-recruitment sites (CSJDM, CSJDC, and HOMO; **Tables S25-S26)**, for a total of n=183 SMI 7:904169:T:TC carriers and n=9,778 SMI non-carriers. Analyses included 75 phenotypes in total: 73 NLP-extracted symptoms distributed across 16 clinical domains (**Table S27**) and two measures of severity (hospitalization status and age at first SMI+ diagnosis). Symptom counts were modeled as rates per visit using zero-inflated negative binomial regression, with sparse features treated as binary outcomes and diagnosis included as a covariate (see **Methods**, **Table S28**); age at first SMI+ diagnosis was modeled using linear regression.

The variant carriers showed markedly higher rates of multiple psychotic, manic, and cognitive/psychomotor symptoms, compared to non-carriers (**Fig. 4**, **Table S29**). Despite the elevated rates of manic and psychotic features, carrier status was not associated with greater overall clinical severity; we observed no difference in rates of hospitalizations or age at first diagnoses between carriers and non-carriers. These findings were also not driven by a specific diagnostic group, by an increased frequency of a severe BD subtype (BD1) among carriers with a BD diagnosis, or by population stratification (**Supplementary Results, Fig. S15-S16, Table S30-S31**).

**Fig. 4:**
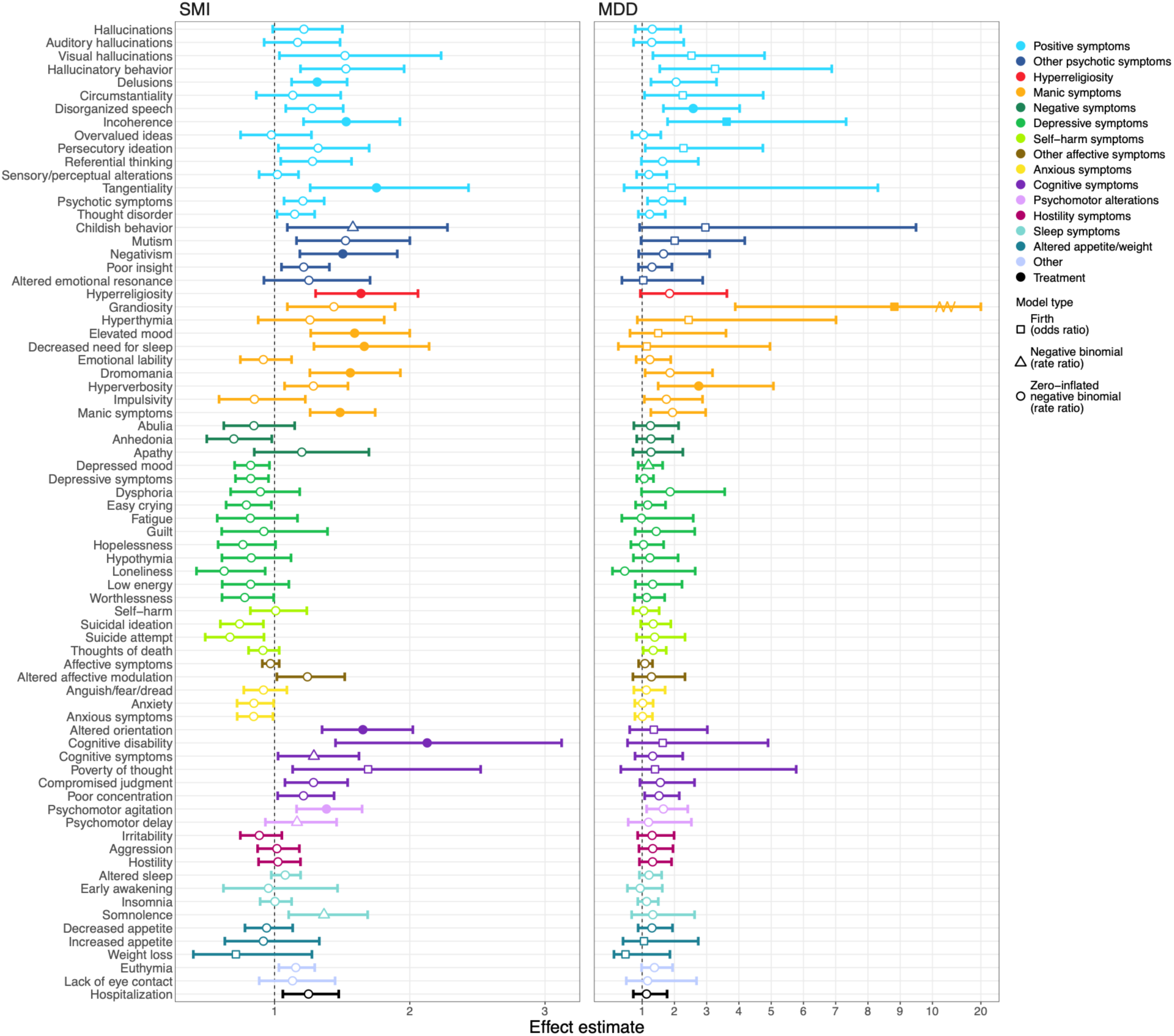
The 7:904169:T:TC variant phenotypically stratifies Mision Origen cases, transdiagnostically. Effect estimates from univariate associations between carrier status and item-level phenotypes shown for participants with SMI (left) and participants with MDD (right), with non-carriers as the reference group. Rate ratios and odds ratios (for phenotypes endorsed by <15% of participants) are shown with bootstrap 95% confidence intervals. Filled points indicate associations surviving Bonferroni correction.

Over-represented psychotic symptoms included positive symptoms that are common features of both SCZ and psychotic forms of BD: delusions (RR=1.32, 95% CI=[1.13, 1.54]; p=6.04e-04), incoherence (RR=1.53, [1.21, 1.93]; p=2.25e-04), tangentiality (RR=1.75, [1.26, 2.43]; p=6.45e-04), and negativism (RR=1.50, [1.19, 1.91], p=6.27e-04, a less common symptom that is often a manifestation of catatonia). Over-represented manic symptoms included: decreased need for sleep (RR=1.66, [1.29, 2.14]; p=4.81e-05), dromomania (an uncontrollable impulse to wander, RR=1.56, [1.26, 1.93]; p=3.04e-05), and elevated mood (RR=1.59, [1.27, 2.00]; p=5.06e-05). Carriers also demonstrated elevated rates of hyperreligiosity, an intense preoccupation with religious or spiritual themes that is particularly characteristic of manic phases of BD (RR=1.64, [1.30, 2.06]; p=1.75e-05). Cognitive and psychomotor symptoms that were more common in carriers compared to non-carriers include psychomotor agitation (RR=1.38, [1.16, 1.65]; p=2.13e-04), altered orientation (RR=1.65, [1.35, 2.02]; p=8.49e-07), and clinician reports of cognitive disability (RR=2.13, [1.45, 3.12]; p=7.44e-05), **Fig. 4**, **Table S29**.

The elevated frequency with which cognitive disability was reported in the EHR of SMI-affected 7:904169:T:TC variant carriers compared to non-carriers motivated us to perform a similar comparative analysis of carriers and non-carriers on scores from cognitive performance assessments (nine modules in the Penn Computerized Neurocognitive Battery; with speed and accuracy components for a total of 17 measurements, Table S26) that we had obtained on a subset of the SMI-affected participants (n=2,398 participants with SMI diagnoses), in previous work^29,45^. In those studies individuals with diagnoses of SCZ and BD1 had demonstrated substantially lower speed and accuracy compared to controls on a broad range of measurements (with particular decrements in executive function [where effect sizes neared and exceeded 1 SD], social cognition, complex cognition accuracy and motor praxis speed tests). Approximately 70% of participants were able to complete all 9 modules (**Table S26**); such completion was not associated with variant carrier status (OR=0.84, p=0.56). One measurement (accuracy of the digit substitution test, an assessment of executive function) showed significantly lower cognition scores in carriers than in non-carriers, after Bonferroni correction (**Fig. S17**), with an effect size of ∼0.50 standard deviation (SD) units. This measurement had shown large deficits (approaching or exceeding 1 SD unit) in BD1 and SCZ individuals relative to controls, in our previous studies^45^. Direct cognitive assessment of a larger number of 7:904169:T:TC carriers will be needed to precisely characterize the impact of this variant on specific cognitive domains and to determine whether the observed deficits are broader than those detected here.

Although the 7:904169:T:TC variant is not significantly associated with MDD, MDD is a highly heterogeneous disorder that shares substantial genetic and clinical overlap with BD^15,16,46,47^, particularly for the severe forms of depression represented in Mision Origen. We therefore examined whether item-level clinical phenotypes could distinguish carriers from non-carriers within the MDD diagnostic group as a means of dissecting this heterogeneity. By carrying out a similar analysis to that used for SMI, we compared n=54 MDD 7:904169:T:TC carriers with MDD to n=7,630 MDD non-carriers. We observed that carriers had a significantly higher frequency of two symptoms typical of BD, grandiosity (OR=8.82, [3.89, 20.02]; p=1.24e-07) and hyperverbosity (RR=2.75, [1.50, 5.07], p=6.67e-04), and two positive symptoms, incoherence (OR=3.62, [1.79, 7.32]; p=2.14e-04) and disorganized speech (RR=2.58, [1.66, 4.02]; p=1.50e-05), **Table S29**.

We considered the possibility that unrecognized BD or schizoaffective disorder could explain the finding of increased rates of BD-like and positive psychotic symptoms in 7:904169:T:TC variant carriers with MDD. To explore this possibility, an experienced research psychiatrist performed a blinded chart review of MDD carriers, together with a closely matched set of MDD non-carriers, assigning a best-estimate diagnosis from lifetime clinical records. From the 52 MDD carriers that were matched using this procedure, the initial MDD diagnosis was confirmed in 49 carriers (94%) and in all matched non-carriers (**Table S32**). The remaining three carriers received a best-estimate diagnosis of BD, including one whose records documented a conversion from MDD to BD after recruitment. Manual symptom annotation further confirmed broadly increased odds of manic and psychotic features in carriers (**Fig. S18, Table S33**), corroborating the findings based on automated NLP-derived phenotyping.

### 7:904169:T:TC disrupts ADAP1 protein structure

As a first step toward elucidating the mechanisms through which the 7:904169:T:TC variant exerts phenotypic effects, we examined its predicted impact on ADAP1 structure. The 7:904169:T:TC frameshift variation seriously compromises the pleckstrin homology domains 1 and 2 (PH1, PH2) located at the C-terminus of the protein (**Fig. 5a**). Wild type ADAP1 acts as an adapter; the PH1 and PH2 domains perform the adapter function, connecting the motor protein KIF13B, to its cargo – a vesicle which contains a specific phospholipid that stimulates neurite outgrowth, phosphatidylinositol (3,4,5)-trisphosphate (PIP3)^48^ (**Fig. 5b-c**). By tethering PIP3-containing vesicles to this motor, ADAP1 enables transportation of PIP3 to the dendrite tip, consequently promoting development of the dendrite into an axon^49^. Specifically, PH2 and a large portion of PH1 are deleted in the variant ADAP1 protein, and replaced with a 5-residue appendage (**Fig. 5d**). One consequence of this truncation might be a loss of ADAP1 function; that is, reduced transport of PIP3 to dendrite tips, leading to altered axon development. An alternative consequence might be a gain of toxic function caused by exposure of an amyloid-prone segment that would otherwise be buried in the center of the PH1 domain (**Fig. 5d**). The program ZipperDB predicts that the segment, 194-GLQVTY-199, has a strong propensity to form a steric zipper, a fundamental structural motif of amyloid fibrils, exhibiting an energetic stability (-26 kcal/mol), that exceeds the threshold score (-23 kcal/mol)^50^. At elevated concentration of the variant ADAP1 protein, it might aggregate into stable amyloid fibrils, as is known to occur in multiple neurodegenerative diseases^51^ (**Fig. 5e**). Amyloid formation would only be favored under the scenario that the truncated transcript was able to escape nonsense mediated RNA decay (NMD), affording the chance for truncated ADAP1 proteins to aggregate; however, we cannot currently predict this possibility, which depends on several variables. Structural evidence strongly implies that truncation of ADAP1 caused by the variant would alter ADAP1 activity, but how this disruption leads to mental illness is not yet clear.

**Fig. 5:**
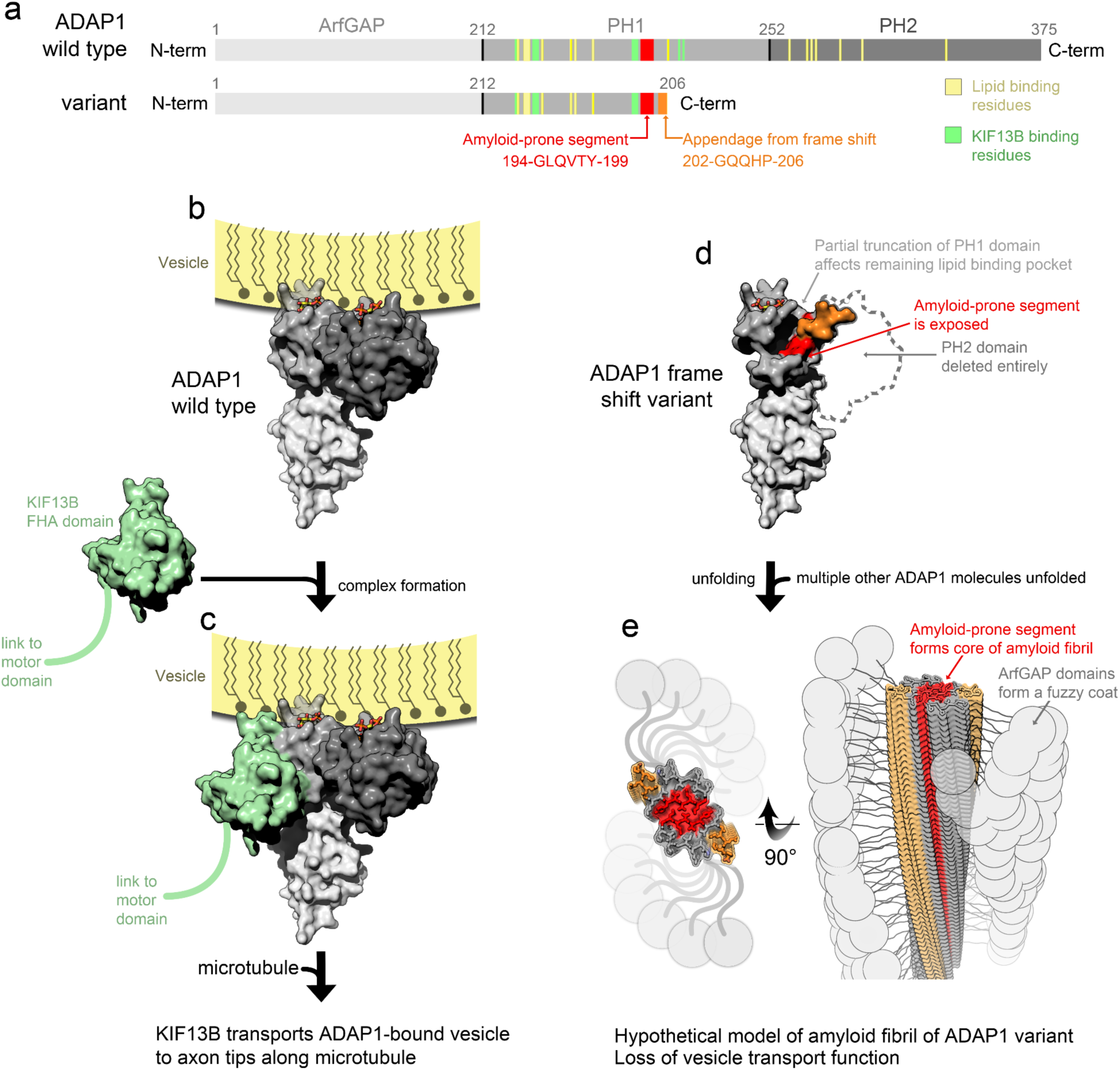
A model for how the frameshift mutation 7:904169 T:TC could affect the structure and function of ADAP1. (a) ADAP1 is composed of three domains: the ArfGAP domain which inactivates a regulator of vesicular trafficking called ARF6 (light gray), and two pleckstrin homology (PH1 and PH2) domains (darker gray) which bind to the polar head group of a phospholipid involved in axonal development, called phosphatidylinositol (3,4,5)-trisphosphate (PIP3). The PIP3-binding residues (yellow) of the two PH domains tether ADAP1 to its cargo vesicle. ADAP1 also binds to the forkhead-associated (FHA) domain of a motor protein called KIF13B, which drives transportation of the ADAP1-tethered vesicle along a microtubule to the axon tip. The KIF13B-binding residues (green) are in PH1. The frameshift mutation replaces PH2 and a large portion of PH1 with a 5-residue appendage (orange). An amyloid-prone segment (red) is present in both the wild type and variant ADAP1. (b) Structure of wild type ADAP1 (PDB ID 3lju) bound to the polar head group of PIP3 (sticks). The cargo vesicle is modeled as a yellow shell near the lipid binding pockets of PH1 and PH2. (c) Structural model of the cargo-loaded ADAP1 bound to the FHA domain of the motor protein KIF13B (PDB ID 3mdb). (d) A structural model of variant ADAP1 illustrates three key points: the loss of PH2 and its lipid binding pocket, compromise of the lipid binding pocket in PH1 resulting from the loss of its C-terminal half, and exposure of the amyloid-prone segment (red). (e) A model of amyloid fibril formation that might result from exposure of the amyloid-prone segment. In this model the amyloid-prone segment of one molecule pairs with an identical segment from another molecule, forming a zipper interaction that leads to fibril formation^90^.

## Methods

### Cohort description

Analyses were conducted using the Mision Origen cohort, an EHR-linked biobank including 91,504 participants, developed for the transdiagnostic study of SMI and severe MDD, including n=44,996 case and n=46,508 control participants. Cases with SMI and severe MDD (n=44,996) were ascertained through electronic medical records of four psychiatric hospitals serving the Paisa region of Colombia: HOMO (in Bello), SAMEIN (in Medellín), CSJDC (in La Ceja), and CSJDM (in Manizales). Case eligibility was based on a primary ICD-10^10^ diagnosis of SCZ [including schizoaffective disorder], delusional disorder (DD), BD, or MDD, as defined by qualifying recruitment diagnoses (**Table S1**).

Within Mision Origen, a subset of 9,107 participants, including 7,686 case participants, was recruited through the Paisa Project^29,45^, which preceded Mision Origen and later became part of the broader biobank. These participants were also ascertained through EHR, but project diagnoses were obtained through the NetSCID, a computerized version of the Structured Clinical Interview for DSM-5)^52^ and additional cognitive assessments were performed using the Penn Computerized Neurocognitive Battery^53^. For these participants, severity/recurrence of MDD was defined by the requirement of at least one of the following: psychiatric hospitalization, treatment for symptoms warranting hospitalization, history of suicide attempt, history of psychosis, or history of electroconvulsive therapy.

Nearly all control participants (n=45,087, 97%) were recruited through five primary care clinics operated by SURA, one of the largest health insurance and healthcare providers in Colombia, with recruitment sites located in the same metropolitan areas represented in case ascertainment. Controls were required to have no current or lifetime history of psychotic or mood spectrum disorders, based on self-report and further EHR review. The Paisa Project additionally contributed 1,421 control participants, ascertained from the same communities as cases, and recruited from friends, neighbors, or in-laws of cases, or from university students and staff, and hospital staff. To participate in the biobank, all participants were further required to reside in the Paisa region, be 18 years of age or older, and have the capacity to provide informed consent. Individuals were excluded if they had significant intellectual disability, a history of a major neurological disorder, or symptoms primarily attributable to substance use. Mision Origen recruitment covered the entire Paisa region: of study participants with recorded birthplace information (n=90,426), almost all (n=84,769, 94%) were born in the Paisa region, covering 178 of its 178 municipalities.

Each participant provided a whole blood sample for DNA extraction and sequencing. The analyses presented here represent the first Mision Origen data freeze, comprising n=52,848 sequenced samples.

### Ethics approval and consent

The Mision Origen and Paisa studies were conducted in accordance with applicable ethical and regulatory requirements and the principles of the Declaration of Helsinki. The study protocol was reviewed and approved by the ethics committees of the Universidad de Antioquia (UdeA), SURA, and each participating psychiatric institution where recruitment was conducted, as well as by the UCLA Medical Institutional Review Board 3 (IRB-20-0149 and IRB-16-2084). All participants provided written informed consent for study participation and for the collection and research use of biological samples, genetic data, clinical information, and information derived from electronic health records. Participant privacy and confidentiality were protected throughout data collection, processing, and analysis.

### EHR data and other study measures

Electronic health records of study participants were available dating back as long ago as 2005, with dates depending on the start of their respective EHR systems (**Table S25**). EHR information leveraged in this study includes longitudinal diagnostic codes (ICD-10), inpatient and outpatient health care utilization, and a broad range of item-level phenotypes extracted from clinical notes^7–9^. As municipality of birth is not systematically recorded in EHRs, it was independently collected for all participants. Cognitive assessments using the Penn Computerized Neurocognitive Battery (CNB) were additionally available for participants with SMI diagnoses within the Paisa Project subset (n=2,398; see **Table S26**).

### Diagnosis assignment and diagnostic phenotypes

Project diagnoses for all participants were recorded at the time of recruitment. For our main analyses, diagnoses were grouped as follows: SMI, comprising SCZ (which includes schizoaffective disorder) and BD; and SMI+, comprising SMI and MDD. We further performed diagnosis specific analyses (SCZ, BD, and MDD). In secondary analyses, we aimed to restrict BD to individuals with a diagnosis of bipolar disorder type I (BDI). As ICD-10 does not distinguish BDI from other types of BD diagnoses, BDI was operationalized as individuals with a project diagnosis of BD combined with either a manic episode recorded in the EHR (F31.1, F31.2, F31.6, F30.1, F30.2) or a NetSCID diagnosis of BDI. Individuals with DD were excluded from analyses presented in this manuscript as were control participants with any eligible case diagnosis recorded in their record (**Fig. S1**).

### Genetic analyses

#### Sequencing generation and variant calling

DNA samples used in BGE-DRAGEN were extracted from whole blood specimens. Following DNA extraction, the BGE protocol was followed, as described by Boltz et al 2026^33^. Joint calling of samples was performed using Illumina DRAGEN^54^. Alignment and variant calling were performed with the DRAGEN Bio-IT Platform, which provides hardware-accelerated algorithms for read mapping, duplicate marking, local realignment, and base quality recalibration within a single pipeline. Variants were called per sample in gVCF mode, capturing both variant and non-variant positions with associated likelihoods. For the DRAGEN callset, BGE-DRAGEN, sample gVCFs were merged and joint genotyping was performed using the Genomic Variant Store (GVS) joint-calling workflow under default best-practice parameters.

#### Exome QC

We used Hail^55^ to write the GVS as a Hail matrix table. Next, we removed sites with more than 6 alternate alleles present, outside of the Twist exome capture target intervals or within low complexity regions. We split multi-allelic sites into biallelic sites and removed variants failing VariantExtractTrainScore (VETS) algorithm, which is the functional equivalent of GATK’s VQSR for DRAGEN data. Genotype-level filters were applied as follows. Homozygous reference genotypes were required to have genotype quality (GQ) ≥ 25. Homozygous alternate genotypes were required to have GQ ≥ 25 and sequencing depth (DP) ≥ 12. Heterozygous genotypes were required to have GQ ≥ 25, DP ≥ 12, and allele balance consistent with heterozygosity, defined as either: (AD_ref_+AD_alt_)/DP<0.8 or AD_alt_/DP>=0.25. Variants that were monomorphic after genotype filtering were removed. Variants with call rate < 0.95 were also excluded. After variant-level quality control, the dataset contained 18,956,828 variants, of which 2,156,906 fell within TWIST regions and were included in the analysis. All quality control procedures were performed using Hail.

#### Sample QC

We performed a series of quality-control checks, summarized in **Fig. S1**. Samples that withdrew consent or failed administrative quality control were removed (n=67). To identify genetic outliers, analyses were restricted to autosomal variants, and several sequencing quality metrics were evaluated. Samples were excluded if they had a chimera rate or contamination greater than 5%. Additional filters were applied to remove samples with an excessive number of singleton variants (>2,500), extreme transition/transversion ratios (Ti/Tv > 3.5 or < 2.5), or extreme heterozygous-to-homozygous variant ratios (>3.5). We then evaluated the distributions of additional sequencing metrics, including the Ti/Tv ratio, the numbers of insertions and deletions, transitions and transversions, the heterozygous-to-homozygous variant ratio, the insertion-to-deletion ratio, the number of singleton variants, and the total number of SNPs. Samples exceeding eight median absolute deviations from the mean for any of these metrics were removed, see **Fig. S2** for distributions. Following these filtering steps, 3,383 samples were excluded. Samples with missing X chromosome data were removed (n = 36). Genetic sex was inferred using the number of variants observed on the Y chromosome; samples with YCOUNT ≤ 1,000 were classified as female and those with YCOUNT > 1,000 as male (see **Fig. S3** for the distribution). Samples with discrepancies between genetic and self-reported sex were removed (n=98), as were duplicated samples (n=57). This analysis was performed in Hail. Following genetic QC, we removed controls without EHR data unless they had been screened through structured clinical interviews by clinicians (n=176) and those with an ICD code corresponding to an SMI diagnosis (n=1,268; **Table S1** lists ICD codes). Related individuals up to second-degree (PI-HAT>=0.25) were removed (n=5,851). When selecting among related individuals, priority was given to samples with available cognitive and behavioral phenotypic data, samples with a diagnosis of SCZ or BD, and samples with available EHR data. Relatedness inference was performed using plink, NaToRA^56^, and PRIMUS^57^, see **Supplementary Text S1** for more details on this procedure. Finally, cases without a diagnosis of MDD, SCZ [including schizoaffective disorder], or BD were removed from the analysis (n=128). The remaining n=41,784 samples were included in the genetic analysis, of which 19,826 were controls, and 21,958 were cases.

#### Imputed data QC

Imputed genotype data were obtained from running GLIMPSE2^58,59^ on the hg38 1KG + HGDP reference panel^60^. Following the process laid out in BGE imputation development^33^, indels and singletons in the reference panel were excluded prior to imputation, resulting in approximately 70.3 million SNPs to be imputed. GLIMPSE2 low-pass WGS imputation was performed using default parameters in two separate runs, comprising n=16,488 and n=37,404 samples, respectively. Within each run, sample batches of 750 individuals were imputed, and all batches merged prior to INFO score calculation and post-imputation QC. Variants with imputation quality scores (INFO) < 0.8 were removed independently in each run. Paisa and Mision Origen imputation runs were subsequently merged, retaining only variants that passed imputation quality control in both batches and filtering to samples that passed exome-based QC (n=41,784 samples). Genotypes with posterior genotype probability < 0.8 were set to missing, and variants with call rate < 0.95 were excluded. Multi-allelic variants were removed using a two-step approach. First, variants with MAF < 0.1% were removed to exclude sites where a single alternate allele occurred at very low frequency in the dataset. The remaining multi-allelic variants were then completely removed, regardless of frequency. Hardy–Weinberg equilibrium filtering was applied in control samples only, excluding variants with p < 1e-15. Finally, the imputed genotype dataset was merged with the exome sequencing dataset, and for variants present in both datasets, the exome sequencing genotypes were preferentially retained. All quality control procedures were performed using Hail.

#### Single-variant association analysis

After sample-level quality control, 1,768,153 non-monomorphic variants remained for analysis. Variants were annotated using Ensembl Variant Effect Predictor (VEP 95)^61^. Variants annotated as transcript ablation, splice acceptor, splice donor, stop-gain, or frameshift were classified as protein-truncating variants (PTVs), and high-confidence loss-of-function variants were identified using the LOFTEE v1 framework. Damaging missense variants were defined using the same approach as the SCHEMA2 and BiPex2 consortium efforts, with the damaging score obtained directly from BiPex2 authors. The score is an ensemble score which included MPC v2, AlphaMissense, and MisFit-S^4,5^. Briefly, a mean-missense Z-score was calculated for each variant: each missense score was ranked across the exome, ranks were converted to Z-scores, and the resulting Z-scores were averaged across methods. Variants with a mean score >=93^rd^ percentile were classified as damaging. After restricting to these deleterious variants, 74,535 variants remained. For single-variant genome-wide association analyses, variants were further filtered to require a MAC ≥ 3 ensuring at least two carriers per variant, yielding 21,506 variants (**Fig. S4**). Association analyses were performed using SAIGE^62^ with Firth logistic regression to account for case–control imbalance, incorporating a genetic relationship matrix (GRM) and adjusting for the first 5 principal components (PCs) of genetic ancestry. PCs were calculated using GCTA (v1.9x)^63^ on imputed data with MAF>5% and LD pruned using PLINK v1.9 (--indep-pairwise 500 50 0.5). Genome-wide significance was defined as p<5.00e-09, as suggested for rare variant studies^64^, and FDR significance was defined at a 5% false positive rate using Benjamini-Hochberg procedure, applied separately for each phenotype (SMI, SMI+, SCZ, BD, MDD): SMI, p ≤ 1.62e-06; SMI+, p ≤ 9.16e-14; SCZ, p ≤ 1.61e-10; BD, p ≤ 1.36e-18; MDD, no p-value met the FDR-corrected significance threshold.

#### Gene-based burden analysis

We performed a gene-based burden test sensitivity analysis, considering both the full set of 74,535 damaging variants and a restricted set limited to variants with MAC<=5, which reduced the total to 60,119 variants. Association analyses were performed using SAIGE-GENE^65^ with Firth logistic regression to account for case–control imbalance, incorporating a GRM, adjusting for the first 5 principal components of genetic ancestry, and applying no weighting by variant frequency. Exome-wide significance was defined using Bonferroni correction at an alpha level of 0.05 for each phenotype, considering all genes tested across both variant filters (no filter and MAC<=5): SMI, p ≤ 2.51e-06; SMI+, p ≤ 2.30e-06; SCZ, p ≤ 2.80e-06; BD, p ≤ 2.63e-06; MDD, p ≤ 2.57e-06. FDR significance was similarly defined at the phenotype level, pooling p-values across both variant filters and applying a 5% false positive rate using Benjamini-Hochberg procedure: SMI, p ≤ 1.38e-05; SMI+, p ≤ 2.83e-06; SCZ, p ≤ 1.05e-09; BD, p ≤ 5.28e-06; MDD, no p-value met the FDR-corrected significance threshold.

#### Common variant conditional analysis

To evaluate common variant associations in the regions harboring the 7:904169:T:TC and 17:68433486:A:G variants, we performed chromosome 7-restricted and chromosome 17-restricted GWAS using SAIGE logistic mixed models, incorporating a genetic relatedness matrix and adjusting for the first five principal components of ancestry. Variants with MAF ≥ 5% were tested for association with SMI and SMI+. Within a previously defined regions of interest containing the 7:904169:T:TC variant (7:1,817,033– 2,283,308 basepairs^29^) and the 17:68433486:A:G variant (17:67,803,484–68,079,772 basepairs^29^), based on published work^1–3,34^, independent common variant signals were identified by LD clumping in PLINK, with LD estimated directly from study participants. Index variants were defined at genome-wide significance (p < 5.00e-08); secondary variants were included at p < 1.00e-05, r² < 0.50, within a 500 kb window. For the 7:904169:T:TC variant, the analysis yielded six independent loci for SMI of which two were shared with SMI+ (**Table S10**). We verified that these lead variants removed the signal in the common variant analysis (**Fig. S7-S8**), and then tested the independence of the 7:904169:T:TC variant association from common-variant signals. Conditioning was performed by including lead SNP dosages as additional covariates in the model, both individually and jointly. Analysis was performed using SAIGE as the primary analysis. As there were no significant associations identified in the previously defined region containing the 17:68433486:A:G variant, conditional analyses were not performed for this locus.

#### Local ancestry inference of 7:904169:T:TC and 17:68433486:A:G carrier haplotypes

To infer local ancestry of the 7:904169:T:TC variant, we restricted quality-controlled genotype data to the first 20 megabases of chromosome 7 (positions 0–20,000,000), the region containing the variant. For the 17:68433486:A:G variant, we restricted genotype data to the 20 megabases surrounding the variant (positions 58,433,486-78,433,486). Allele frequencies were computed across both regions, and variants with MAF < 0.01 and strand ambiguous AT/GC variants were excluded from downstream analyses. The 7:904169:T:TC variant, which has a MAF < 0.01, was retained separately and merged onto its corresponding dataset. Haplotype phasing of the two subsets was then performed using Eagle v2.4.1^66,67^. We inferred three-way local ancestry using RFMix v2.03-r0^41^. Reference population data were obtained from the Human Genetic Diversity Project (HGDP) and 1000 Genomes Project (1kGP)^60^, and subset to the following populations: AMR, (n=549), EUR (n=752), and African (AFR, n=992). As the AMR samples in HGDP and 1kGP are recently admixed, we excluded 358 AMR samples from the reference with < 90% Amerindigenous ancestry, following the methods described by Boltz et al.^33^ The resulting reference population included 1935 individuals (AFR n=992, AMR n=191, EUR n=752).

#### Local ancestry-informed association analysis

To conduct local ancestry-informed association analyses, we employed Tractor-Mix^42^, a generalized linear mixed model (GLMM) extension of the original Tractor framework^68^. Associations were examined for the 7:904169:T:TC variant, significant variants from conditional analyses (Table S6), and the 17:68433486:A:G variant, across phenotypes including SMI, SMI+, SCZ, BD, and MDD. Ancestry-specific allele dosages and haplotype counts were extracted from RFMix output using the Tractor extract_tracts function for all three ancestries (EUR, AFR, and AMR). All associations included a sparse GRM and the first five genetic PCs as covariates. The allele count threshold was set as 50, except for 17:68433486:A:G. As there are fewer carriers with this variant, the allele count threshold was set to 10, lower than the number of carriers with MDD.

Both unconditional and conditional analyses were performed. Conditional analyses included ancestry-specific haplotype counts to condition on local ancestry dosage at each tested locus and were only performed for variants with an allele count greater than 50 for all three ancestries.

#### Comparing relatedness in variant carriers and non-carriers

We used PLINK to contrast identity-by-state (IBS, a measure of genetic distance) between pairs of carriers and pairs of non-carriers, separately for 7:904169:T:TC and 17:68433486:A:G, using the --ibs-test command. P-values were estimated using 1,000,000 permutations.

#### Gene expression

Single-nucleus expression was taken from Siletti et al.^44^ and Wamsley et al.^43^ Cell types were harmonized to BICCN subclass nomenclature for cortical subtypes. Cell types represented by fewer than 20 were excluded.

#### Developmental expression

Developmental expression was assessed using the BrainSpan developmental transcriptome as conditional quantile normalized (CQN) gene-level expression across 393 samples from 36 donors. Analysis was restricted to the 11 cortical structures.

#### Protein–protein interaction networks

Interactors of ADAP1 and WIPI1 were retrieved from BioGRID (REST API, Homo sapiens, taxon 9606, version 5.0.259) and STRING (version 12) restricted to the physical subnetwork and combined as a union: a gene was retained as an interactor if reported by either source. Interactions among the resulting gene sets were then queried from both databases and combined as a union, such that an edge was drawn if reported by either database. STRING edges were filtered at a combined score of at least 0.7 (high confidence). Partners retaining no edge after filtering were omitted from plots. Networks were laid out using a force-directed (Fruchterman–Reingold) algorithm, with node size scaled by degree. Over-representation analysis was performed on each interactor set, including the hub gene, using a hypergeometric test (fgsea::fora version 1.24) against Gene Ontology biological process gene sets from MSigDB collection C5 (msigdbr version 7.5.1). Gene sets containing between 5 and 2000 genes were tested, and the background comprised all genes annotated in the tested gene sets. P-values were adjusted for multiple testing using the Benjamini– Hochberg method, and the five most significant terms per gene set are reported.

#### Geographic location of 7:904169T:TC and 17:68433486:A:G variant carriers

To assess whether birthplaces of 7:904169:T:TC and 17:68433486:A:G carriers were geographically clustered relative to non-carriers, we used Moran’s I Global Statistic of spatial autocorrelation^69^, restricted to participants with recorded municipality of birth. For each municipality with at least one recruited participant, we calculated the proportion of carriers of each variant relative to all participants recruited there. Significance was estimated using 1,000,000 Monte Carlo simulations (see **Supplementary Methods** for sensitivity analyses and further details).

To visualize the geographic distribution of 7:904169:T:TC carriers and a randomly selected subset of non-carrier participants of equal sample size, each individual was assigned a location by applying random jitter around the city center of their municipality of birth. Municipal boundary shapefiles were obtained from the geoportal of the Departamento Administrativo Nacional de Estadística (DANE)^70^.

#### Item-level phenotypic analyses

To delineate the impact of carrier status on clinical presentation, we tested whether item-level phenotypes extracted from the free text of clinical notes were associated with carrier status in individuals with the same diagnoses. In addition to EHR-derived phenotypes, we evaluated differences in cognitive performance across domains in the subset of participants with an SMI diagnosis and available data from the Paisa Project (**Table S26**).

#### Item-level phenotypes extracted from clinical notes

From the free text of clinical notes, we extracted a broad set of item-level measures spanning psychiatric symptoms, behaviors, exposures, and treatment characteristics. For the analyses presented here, we focused on clinical symptoms and behaviors directly relevant to psychiatric presentation. Item-level phenotypes were extracted using two complementary natural language processing (NLP) approaches developed and validated on these notes: a traditional pattern-based method^7^, and large language model (LLM)-based extraction^8,9^. Only phenotypes that could be reliably extracted, achieving an F1 score greater than 0.8 (a metric balancing precision and recall), were included. When a phenotype could be reliably extracted using the pattern-based method, that approach was selected for computational efficiency; the remaining phenotypes were extracted using LLMs. Analyses included 75 phenotypes in total: 73 NLP-extracted symptoms and two measures of severity [hospitalization status and age at first SMI+ diagnosis] (**Table S27**). To aid interpretation, phenotypes were grouped into symptom domains based on grouping described in Correll et al.^71^, with a few additional domains added to accommodate features that did not fit into their original scheme: psychomotor alterations, altered appetite/weight, other psychotic symptoms, religiosity, other affective symptoms, treatment, and other, resulting in 16 domains.

#### Univariate associations between 7:904169T:TC carrier status and item-level phenotypes

Participants with complete visit information for at least one inpatient hospitalization or outpatient visit and complete genetic information at the 7:904169T:TC variant site were included in analyses (**Table S25**).

All analyses were conducted separately in participants with SMI and in participants with MDD, as baseline phenotype prevalence differed across these groups. All models described below were adjusted for age at recruitment, sex, recruitment site, and diagnosis (for SMI analysis), in addition to model-specific covariates. Confidence intervals and two-sided p-values were calculated based on distributions of 1,200 bootstrap iterations (see **Supplementary Methods**). A Bonferroni threshold was applied to account for multiple testing (p < 0.05/75 phenotypes).

For each NLP-extracted item-level phenotype, the specific model used for analyses depended on the feature distribution of the item. Common items (> 15% of participants having documented mentions of the item) were modeled as counts, representing the total number of documented occurrences in a patient’s EHRs (recorded at most once per visit), as count-based measures may better capture clinical severity. To account for overdispersion, either negative binomial (NB)^72^ or zero-inflated negative binomial (ZINB) regression^73,74^ was used (**Fig. S19-S20**). For each analysis, model selection between NB and ZINB was determined by the likelihood ratio test (**Table S28**). Total visit count was included as an offset to model phenotype counts as rate per total visits. In ZINB models, total visit count was also included as the logit component to account for the possibility that absent phenotype recordings reflect limited documentation opportunities. Phenotypes that were rare (≥ 85% of participants having no documented mentions of the item) were modeled as binary outcomes using Firth logistic regression instead of counts (**Table S28**). The logistic models also adjusted for total visit count in addition to previously mentioned covariates. The effect of hospitalizations was modeled using the same approach as the NLP-extracted phenotypes. The effect of carrier status on age at first SMI+ diagnosis was modeled using linear regression, restricted to participants with a recorded ICD-10 diagnosis of SMI+.

As a sensitivity analysis, we conducted the same univariate associations between carrier status and item-level phenotypes separately in participants with BD, BDI, SCZ, and in participants with SCZ + BDI. We additionally conducted a sensitivity analysis incorporating the first five genetic PCs into the item-level phenotypic analyses as covariates to adjust for potential population stratification.

#### Association between 7:904169:T:TC carrier status and cognition in variant carriers and non-carriers

Cognitive performance was measured in a subset of participants originally recruited as part of the Paisa Project^29^ using the Penn Computerized Neurocognitive Battery (CNB)^53,75–77^. The CNB measures executive functions (abstraction and mental flexibility, sustained attention, working memory), episodic memory (verbal, facial, spatial, associative), complex cognition (language reasoning, nonverbal reasoning, spatial processing), social cognition (emotion identification, emotion intensity differentiation, age differentiation), and psychomotor speed. The seventeen measures employed in our work are listed in Table S26 along with the number of carriers with cognitive data for each measurement. In the full cohort, cognition was regressed on age, age^2^, age^3^, sex, maximum education level, and the interaction between age and education, and the residuals from this regression were quantile-normal transformed for further analysis. Data for speed of cognition were multiplied by -1 so that poorer performance (longer response time) would result in a lower value. Residual cognition was contrasted between 7:904169:T:TC variant carriers and non-carriers. As SMI affects cognitive performance, we restricted our comparison to carriers/non-carriers with a diagnosis of BD or SCZ, as these diagnoses have similar cognitive deficits^45^. Results were adjusted for multiple testing using a Bonferroni correction for 17 tests.

#### Analysis of chart review in participants with MDD

Each MDD carrier of 7:904169:T:TC was matched to a non-carrier with MDD based on sex, recruitment site, inpatient status (ever hospitalized vs. never hospitalized), age at recruitment, and year of first documented MDD ICD-10 diagnostic code in the EHR. Prior to matching, participants without complete inpatient status or year of first MDD code in the EHR were excluded. Matching without replacement was implemented using the MatchIt package in R^78^, with sex, recruitment site, inpatient status, and year of first MDD code treated as exact constraints and then matched on closest age. An experienced research psychiatrist performed a chart review of all selected MDD participants, blinded to carrier status, recording best-estimate diagnoses and item-level phenotypes based on lifetime clinical records using a checklist adopted from Fears et al.^79^. Differences in item-level frequencies between carriers and non-carriers were assessed using chi-squared test, excluding phenotypes with no documented occurrences in either group.

#### Predicted structure of mutated ADAP1

The amyloid-forming propensities of ADAP1 and the 7:904169:T:TC frameshift variant were evaluated by submitting their amino acid sequences to ZipperDB^50^. The effect of the frameshift variant was evaluated by deleting coordinates from the crystal structure of the wild type ADAP1 protein in complex with KIF13B (PDB ID 3mdb) that correspond to residues following the variant-introduced stop codon^48^. The atomic coordinates were visualized using Pymol (The PyMOL Molecular Graphics System, Version 3.1, Schrödinger, LLC).

## Discussion

In the Mision Origen biobank, we identified five genes with significant impact on SMI or SMI+ that have not previously been implicated in any mental health disorder (*ADAP1*, *WIPI1*, *NFE2L1*, *FBXO10*, and *HLA-DPB1*); the associations in the first four of these genes are driven by individual variants with intermediate frequency in the Paisa population. We also report significant associations to two genes that have previously been highlighted in consortium studies (*KDM5B* and *ZMYM2*). The evidence at two of the novel genes, *ADAP1* and *WIPI1*, exceeds exome-wide significance; for *ADAP1*, a single frameshift variant achieves genome-wide significance for multiple SMI phenotypes.

It has long been hypothesized that intermediate frequency, intermediate penetrance variants are responsible for much of the genetic contribution to common disorders, and that discovering them would have great scientific and medical value, both for elucidating disease mechanisms and selecting targets for drug development^80,81^. At least for major psychiatric disorders, however, these discoveries have not yet occurred. To our knowledge, the findings reported here represent the first step in filling the gap between the two poles of ultra-rare variants identified by the SCHEMA2 and BipEx2 consortia and the common variants of GWAS meta-analyses.

This gap has remained unfilled because, based on expectations for the effects of selection on different types of deleterious variants, discovery of intermediate-frequency large-effect variants requires sample sizes that have not yet been attained for sequencing studies of SMI disorders, even by SCHEMA2 and BipEx2. As modeled by Zuk et al.^19^, the process of rapid expansion in population isolates alters this expectation for any variant that has passed through the founding bottleneck, and therefore the associations to such variants may be detected with a much smaller sample size. Two additional features of the Paisa population accentuate this advantage in comparison with other isolates. Because of the recency of its rapid expansion, selection has had less opportunity to decrease the frequency of deleterious alleles. Furthermore, the large size of the population made it relatively straightforward to recruit the sample of more than 12,000 SMI cases (or almost 22,000 considering SMI+) sequenced in this study.

The founding history of the Paisa may also make it an unusual case among population isolates. The phenomenon of increased frequency of deleterious variants in a rapidly expanded isolate has mostly been observed in European populations and attributed to the effects of random genetic drift. In the Paisa, where this expansion followed early admixture between the indigenous population and European and African immigrants, many of the variants that passed through its founding bottleneck were brought to Colombia by these immigrants, where they faced different selection pressures from infectious diseases than in their continent of origin. It may be important to consider the possible role of balancing selection in increasing the frequency in Mision Origen of variants that we highlight here^82^.

While the dramatically higher frequencies of these variants in the Paisa population reflect the unique demographic history of this founder population, it also highlights the continued underrepresentation of admixed populations generally, and Latin American populations specifically, in global genomic resources. For example, gnomAD v4.1.1 contains only 30,019 admixed American individuals, making it poorly suited to reliably compare allele frequencies of rare variants. Notably, even the Iberian populations that were the primary source of European ancestry in the Paisa are not systematically distinguished in public databases, making it difficult to determine whether the absence of these variants from European databases reflects their true rarity in Europe or gaps in sampling of Iberian populations. Mision Origen is only one of several emerging biobanks constructed in populations that have been underrepresented in genomic databases^83,84^ and that have characteristics favorable for discovering large-effect variants that would be difficult to detect in other genetic research resources. An important objective for the human genetics community should be to rapidly incorporate sequence data from these emerging datasets within the most widely used reference databases, so that all researchers have access to a more complete picture of human allele frequency distributions.

The finding that single variants exert large, nearly equal effects on SCZ and BD provides strong and direct evidence that shared genetic etiology underlies the points of continuity observed across SMI diagnoses – including familial aggregation^85^ patterns, cross-disorder common variant associations and polygenic risk^15,29^, and genes in which the aggregation of ultra-rare variants are associated with both SMI diagnoses^4,5^. As our case sample was recruited from standard treatment settings, without selection for family history or unusual phenotypes, these results likely have generalized importance at the population level. This inference suggests an opportunity, in Mision Origen and the larger Paisa population, to extend our findings to advance two related objectives: stratifying SMI based on causation and identifying the biological basis of this stratification through mechanistic studies of causative variants.

Information routinely available in Paisa region EHRs enabled initial stratification of SMI based on causal factors (carrier status for *ADAP1* variant 7:904169:T:TC) rather than diagnosis; carriers differ significantly from others with SMI for a subset of symptoms typical of psychotic mania (e.g., hyperreligiosity), but we did not detect significant differences for numerous symptoms common in psychotic disorders (e.g., auditory hallucinations) or severe mood disorders (e.g., suicidality or depressive symptoms). Cognitive disability is the feature that, in these EHR analyses, most strongly distinguishes SMI variant carriers from non-carriers. Such disability may reflect impairment in specific dimensions of cognitive performance, as evidenced by the independent observation of substantial deficits in executive function among SMI-affected variant carriers compared to similarly affected non-carriers. In our previous work we showed large deficits on these and other performance measures in SCZ and BD1 participants compared to controls^45^; our current findings suggest that individuals carrying the 7:904169:T:TC variant experience an impairment in cognitive performance on top of that associated with SMI overall.

The results of these initial phenotyping analyses also suggest the potential for more comprehensively stratifying SMI in the Paisa population through prospective multimodal phenotyping of carriers and non-carriers. Given the expected number of carriers, it may be possible to include in these studies assessments of molecular markers (e.g., proteomics) that could inform efforts to identify biological mechanisms. Such phenotyping will also enable stratification of mental illness beyond SMI, as evidenced by our finding that, among individuals with MDD, 7:904169:T:TC carriers display overrepresentation of BD-like features (e.g., grandiosity); further investigations with these individuals may enable prediction of conversion of MDD to BD, a key goal in precision mental health.

Finding single variants that have a strong effect on SMI enables a narrow focus to efforts to leverage genetic results to elucidate disease mechanisms. As we do not yet know the function of a large proportion of the genes that are strongly expressed in the brain, it is not surprising that most of our findings are to genes that have been relatively little studied and not previously linked to these disorders. Notably, our ability to suggest how the 7:904169:T:TC frameshift variation may act to increase risk for SMI is limited by the relative sparsity of existing information about the function of *ADAP1* or its role in disease. In the brain, both ADAP1 and WIPI1 show broad expression across cell types that increases across development. ADAP1 protein interactors point towards its’ role in mediating intracellular vesicular transport while WIPI1 is an effector of autophagy. *ADAP1* also appears to have a role outside of the central nervous system, notably, a gain-of-function screen identified it as an amplifier of T cell signaling through the KRAS–ERK– AP-1 axis and a modulator of latent HIV-1 reactivation^86^, suggesting that the consequences of its truncation may extend to immune function. It has been suggested but not confirmed^40^ that *ADAP1* may play a role in autism spectrum disorders and developmental delay, as it was one of > 250 genes to display an excess of *de novo* missense mutations in a screen of about 10,000 cases^87^.

The 7:904169:T:TC variant is predicted to truncate the domains of ADAP1 (part of PH1 and all of PH2) that perform the protein’s adaptor function. It is possible that, due to NMD, this truncation leads to loss of function such that transport of PIP3 to dendrite tips is reduced, resulting in altered axon development. An alternative consequence of this truncation could be a gain of toxic function caused by exposure of an amyloid-prone segment that would otherwise be buried in the center of the PH1 domain structure. Although we currently have no direct evidence to support either of the above possibilities, studies that generated two different frameshift mutations in the zebrafish orthologue (*adap1*) may be revealing. These frameshifts generate mutants that lack both PH1 and PH2 domains, but that exhibit no obvious phenotypes until adulthood, at which point both heterozygous and homozygous mutants display abnormalities in social and exploratory behaviors^88^.

In conclusion, this study identifies genetic variants contributing to the causation of both SCZ and BD, in genes not previously implicated in mental disorders, and demonstrates that such variation phenotypically stratifies these diagnoses as well as MDD. Further investigations focused on variants carried by up to several hundred participants in the Mision Origen biobank have the potential to identify and validate clinically actionable genetic risk profiles and biomarkers that can be used to predict patient trajectories, match patients to existing treatments, and develop and test new treatments targeting specific mechanisms of psychopathology in subsets of patients.

## Data Availability

Summary statistics are contained in the manuscript. All individual-level data used in this study will be made available through the NIMH Data Archive (NDA) or equivalent repositories, in accordance with data sharing agreements and participant consent.

## Acknowledgements and funding

This study was supported by National Institute of Mental Health grants R01MH113078 (to NBF, CEB, and CLJ), R01MH123157 (to NBF, LMOL, and CLJ), U01MH125042 (to NBF, LMOL, and CLJ), U01MH125047 (to BMN and ARM), R01MH101244 (to BMN), and U24MH140953 (to BMN, LMOL, ARM). UCLA CTSI Grant UL1TR001881 provided support for the data collection system REDCap.This research was funded in part by BD2: Breakthrough Discoveries for thriving with Bipolar Disorder (to NBF, LMOL, CLJ, BMN and JMS) and the Stanley Center for Psychiatric Research at Broad Institute.

## Data availability

Summary statistics are available as part of the supplementary materials. All individual-level data used in this study will be made available through the NIMH Data Archive (NDA) or equivalent repositories, in accordance with data sharing agreements and participant consent.

