## Supplementary Information for "A founder population reveals genetic variation with major effect on serious mental illness"

### Mision Origen Supplementary Methods, Figures, and Tables

#### Supplementary Results

##### Common variant conditional analysis of 7:904169:T:TC

To assess whether the associations 7:904169:T:TC in ADAP1 was independent from associations to common variants previously implicated in SMI or SMI+, we evaluated recent GWAS for BD, SCZ or MDD[1–4] (see **Methods** and Lopera-Maya et al.[5] for details). The closest common variant associations were to BD, SCZ, and MDD, in a region located between 7:1,817,033–2,283,308 basepairs). Additionally, a recent cross-disorder analysis mapping the genetic architecture of 14 psychiatric disorders identified a strong and specific association in this region to a combined SCZ-BD factor[6]. In Mision Origen several loci in this region were significantly associated with SMI and SMI+. In conditional analyses adjusting for the lead SNPs from these associations (**see Methods**), the association with 7:904169:T:TC remained strong and significant (**Fig. S7-S8**, **Table S9**). We additionally identified seven novel associations to common variants (MAF 1-4%) at 7:394,997–1,805,732 that were genome-wide significant and were in weak LD with 7:904169:T:TC (R2 ranging from 0.08 to 0.36; **Fig. S8**, **Table S10**). Conditional analysis confirmed that the effect of 7:904169:T:TC remained consistent after accounting for these variants (**Tables S9-S10**). Conversely, these seven common variants became non-significant when conditioning on 7:904169:T:TC (**Table S10**).

##### Local ancestry of the 7:904169:T:TC and 17:68433486:A:G variants

To confirm that the association signal at 7:904169:T:TC is driven by the EUR ancestry and to estimate ancestry-specific effect sizes, we performed local ancestry-informed association analysis using Tractor-Mix[7], a method that stratifies association tests by local ancestry, enabling ancestry-specific effect estimation in admixed individuals. The association was indeed entirely attributable to EUR ancestry haplotypes across all phenotypes tested: SMI (OR=3.37, 95% CI=[2.64, 4.31], p=2.41e-22), SMI+ (OR=2.30, 95% CI=[1.86, 2.86], p=3.15e-14), SCZ (OR=6.41, 95% CI=[3.90, 10.53], p=2.45e-13), BD (OR=3.58, 95% CI=[2.72, 4.70], p=4.52e-20) and MDD (OR=1.41, 95% CI=[0.99, 1.98], p=5.27e-02), with confidence intervals of effect sizes overlapping those obtained through ancestry-agnostic analysis (**Table S23**).

Similarly, the associations for 17:68433486:A:G were attributable entirely to EUR ancestry, as the variant was present only on EUR ancestry haplotypes: SMI (OR=4.35, 95% CI=[2.42,7.80], p=8.21e-07), SMI+ (OR=2.63, 95% CI=[1.59, 4.36], p=1.81e-04), SCZ (OR=9.81, 95% CI=[2.91, 33.08], p=2.32e-04), BD (OR=4.66, 95% CI=[2.38, 9.14], p=7.45e-06) and MDD (OR=1.98, 95% CI=[0.87, 4.49], p=1.01e-01), with confidence intervals of effect sizes overlapping those obtained through ancestry-agnostic analysis (**Table S23**).

##### Geographic and genetic distance of 7:904169:T:TC and 17:68433486:A:G variant carriers

To test whether carriers share a common geographic origin and excess genetic relatedness we examined spatial and genomic clustering of 7:904169:T:TC and 17:68433486:A:G carriers relative to non-carriers. Of the 7:904169:T:TC variant carriers with recorded birthplace information (n=331), 96% (n=316) were born in the Paisa region. The two metropolitan areas of Medellín and Manizales contribute 58 and 55 carriers, respectively. Carriers showed significant geographic clustering relative to non-carriers at the municipality level (Moran's I=0.08, p=1.75e-02, based on 1,000,000 Monte Carlo simulations), an effect that was consistent when restricting to cases only (N carriers=249, N non-carriers=20,712, I=0.12, p=6.60e-03) and to individuals born in the Paisa region (N carriers=316, N non-carriers =38,067, I=0.15, p=4.75e-03). As expected, 7:904169:T:TC variant carriers were also significantly more genetically related to one another than non-carrier pairs, despite removal of closely related individuals from all analyses: 10.8% of carrier pairs were related at levels between second and first cousin, while only 1% of non-carrier pairs were related at this level. Using PLINK's Identity-by-state testing procedure, in 1 million permutations, we never saw the mean IBS values between non-carrier pairs exceed the mean IBS values between carrier pairs (p<1e-6).

In conducting similar geospatial analyses for 17:68433486:A:G carriers (n=56) and non-carriers (n=39,919) with recorded birthplace information (**Fig. S14**), we also observe modest but significant geographic clustering (I=0.06, p=3.69e-02). This effect was consistent when restricting to cases only (N carriers=45, N non-carriers=20,917, I=0.05, p=5.00e-02) and to individuals born in the Paisa region (N carriers=54, N non-carriers =38,336, I=0.07, p=4.28e-02). Carriers of the 17:68433486:A:G variant were significantly more related to each other than non-carrier pairs, but not to the degree seen for the 7:904169:T:TC variant; 3% of carrier pairs were related at levels between second and first cousin, compared to 1% of non-carrier pairs. PLINK's Identity-by-state testing procedure resulted in a permutation p-value of 2.58e-02 for the hypothesis of equal relatedness in carrier and non-carrier pairs.

##### Sensitivity analyses confirm transdiagnostic item-level phenotypes are associated with 7:904169:T:TC carrier status among participants with SMI diagnoses

To investigate whether elevated rates of manic and psychotic features in 7:904169:T:TC carriers were driven by a specific diagnostic group, even after adjusting for diagnosis as a covariate, we performed the same univariate associations for the Bonferroni significant item-level phenotypes in four diagnostic subsets of SMI: SCZ and BDI together; and SCZ, BD, and BDI, individually. The resulting effect estimates for all diagnostic subsets had overlapping confidence intervals with the original item-level phenotype associations in participants with SMI (**Fig. S15, Table S30**). Additionally, to assess whether the item-level phenotypic findings could be affected by population stratification, we repeated the same item-level phenotypic analyses in participants with SMI and MDD, including the first five genetic PCs as covariates. The resulting analyses also had overlapping confidence intervals with the original results (**Fig. S16, Table S31**).

#### Supplementary Methods

##### Procedure for relatedness filtering

To construct a maximally unrelated set of participants for downstream genetic analyses, we implemented a multi-step relatedness-filtering procedure that incorporated genotype-based relatedness estimates and phenotypic information to prioritize participants with more extensive phenotypic data. A PLINK genome file was generated from all WES variants with MAF>1%, LD-pruned with R2=0.5 and a 500k window, and containing pairwise relatedness estimates. A relatedness threshold of PI_HAT=0.25 was used, corresponding to first-degree relationships including half-siblings, avuncular relationships, grandparent–grandchild relationships, full siblings, and parent–offspring pairs. The relationship-pruning algorithm

NaToRA[8] was applied to identify groups of related individuals ("families") using a kinship threshold of 0.125 or higher (corresponding to PI_HAT=0.25). Among these participants, those unrelated to any other participant at the specified relatedness threshold, as well as those whose only relatives had already been excluded in the previous filtering step, were retained directly for analysis. This group comprised 35,960 participants. The remaining 11,837 participants were related to at least one other individual at the specified threshold. For these participants, we extracted all relevant pairs from the PLINK genome file and assigned family identifiers based on the NaToRA output. A phenotype file was then constructed for these participants containing family identifier (FID), individual identifier (IID), and a phenotype prioritization score reflecting the availability of key phenotypic information. The phenotype score was defined as: HasCNB × 4 + SMI × 2 + EHR, thereby prioritizing individuals with cognitive and behavioral data (HasCNB is an indicator variable for having data from the Penn Cognitive Battery), followed by individuals with SMI (an indicator for the diagnosis of BD or SCZ), and then individuals with electronic health record data (EHR is an indicator for availability of EHR data). The subsetted genome file and the phenotype file were provided as input to PRIMUS[9]. Using the specified relatedness threshold, PRIMUS identified the maximally unrelated set of individuals that also maximized the average phenotype score within each related family cluster. Of the 11,837 related participants, PRIMUS selected 5,986 individuals for retention. Of the individuals removed, 2,952 had a first-degree relationship (of which 1,025 had also a second-degree relationship), and 2,899 had a second-degree relationship. Given the phenotype score, the algorithm favored the removal of controls; of the removed individuals, 3,416 (59%) were controls, 1,425 (24%) had a MDD diagnosis, 766 (13%) had a BD diagnosis, 153 (3%) had a SCZ diagnosis, and 72 (1%) had a schizoaffective disorder diagnosis. Of these excluded individuals, 72 were carriers of the 7:904169:T:TC variant, 28 were controls, and 44 had an SMI diagnosis.

#### Comparison of Mision Origen findings to other biobanks

We compared minor allele frequencies and minor allele counts for burden associated variants across multiple external data resources including the Regeneron Million Exome Variant Browser (RGC), All of Us (AoU), UK Biobank, and the BRAVO variant browser for TopMed Freeze 10. RGC consists of 821,979 individuals with WES, 54,629 (6.6%) of which are designated as Indigenous American Ancestry (IAM). The UK Biobank Allele frequency browser records data for 490,640 individuals with WGS, but notably does not identify AMR/IAM individuals. TopMed Freeze 10, as queried by the BRAVO variant browser, consists of 150,899 individuals with WGS data, of whom ~19% are assigned AMR genetic ancestry. The AoU SNV/Indel browser records data for 535,680 individuals with WGS. AoU has historically reported ~6.3% of individuals with WGS assigned to AMR genetic ancestry (The All of Us Research Program Genomics Investigators, Nature 2024), though this proportion was estimated in a subset (n=245,388) of the AoU WGS sample. Allele frequencies were compared to both EUR and AMR/IAM references where available.

#### Geographic clustering of 7:904169T:TC and 17:68433486:A:G variant

To assess whether birthplaces of 7:904169:T:TC and 17:68433486:A:G carriers were geographically clustered relative to non-carriers, we used Moran's I Global Statistic of spatial autocorrelation[10], restricted to participants with recorded municipality of birth. Moran's I ranges from -1 (perfectly dispersed) to +1 (perfectly clustered), with values near 0 indicating random geographic distribution. For each municipality with at least one recruited participant, we calculated the proportion of carriers of each variant relative to all participants recruited there. Spatial weights were computed from municipality polygon adjacencies using queen contiguity, in which two municipalities were considered neighbors if they shared at least one boundary point. Weights were row-standardized so that each municipality's neighbor weights summed to one. Significance was assessed using 1,000,000 Monte Carlo simulations in which carrier proportions were randomly permuted across municipalities; the p-value was defined as the proportion of permuted Moran's I values greater than or equal to the observed Moran's I.

Two sensitivity analyses were performed: one restricting to case participants only, to account for potential bias arising from differential recruitment locations between cases and controls and the higher carrier frequency among cases; and one restricting the analyses to only include individuals born in the Paisa region (departments Antioquia, Caldas, Risaralda, and Quindío).

#### Item-level phenotypic analyses

To calculate p-values and confidence intervals for each item-level phenotype, we used the following procedure. For each bootstrap iteration, the data were sampled with replacement, and the model was refit, and the new coefficient estimates for each predictor were saved. Repeating this process 1,200 times produced an empirical distribution of coefficient estimates. From this bootstrap distribution, 95% confidence intervals were constructed using a normal approximation, with the original (non-resampled) coefficient estimate as the center of the interval and the standard deviation of the bootstrap replicates used as the standard error. Bootstrap p-values were calculated based on a bias-corrected z-statistic, which was used to compute a two-sided p-value using the standard normal distribution.

#### Supplementary Figures

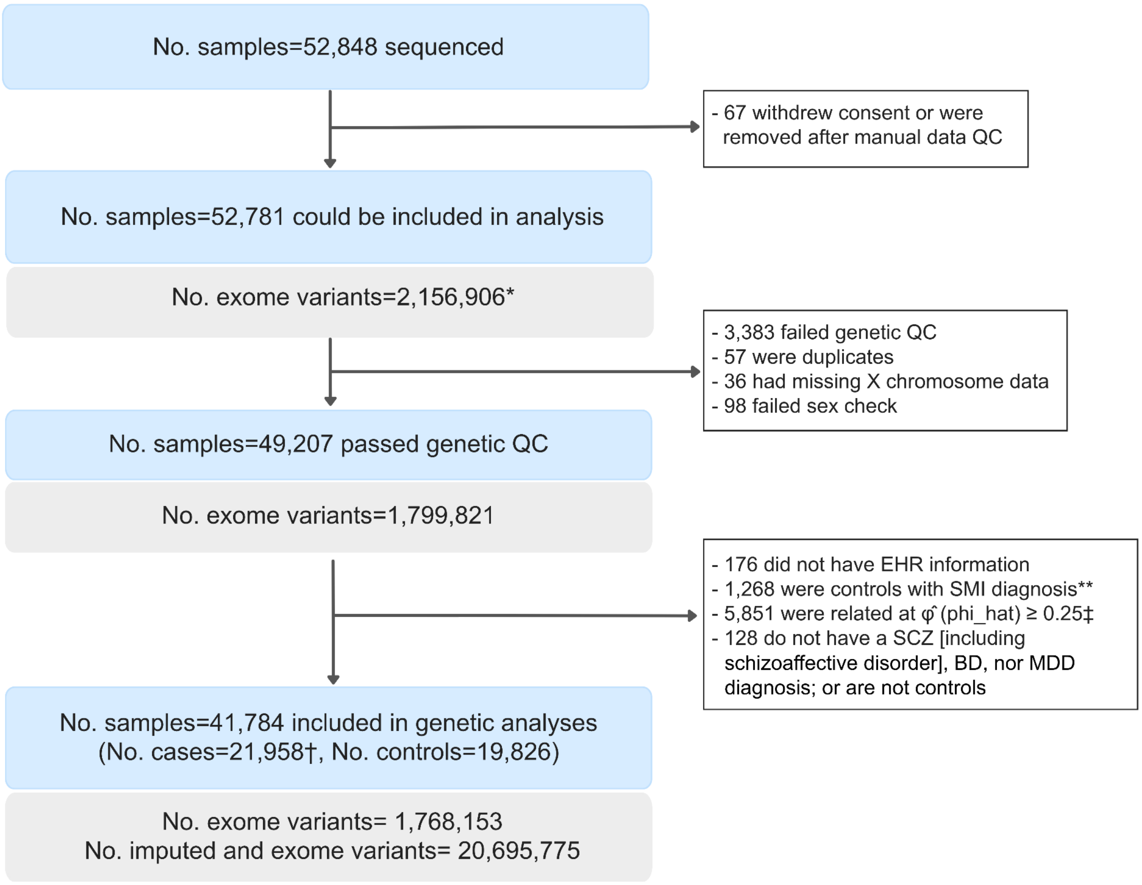

* Number of variants after high-quality filtering and in TWIST regions
† 9,763 Major Depressive Disorder, 8,507 Bipolar Disorder, 3,065 Schizophrenia, 623 Schizoaffective Disorder
‡ 2,919 with a first degree relative (of which 991 had both first and second degree relatives), 2,913 with a second degree relative
** Number of controls with an SMI diagnosis: 1,014 Major Depressive Disorder, 176 Bipolar Disorder, 66 Schizophrenia, 7 Schizoaffective, and 5 Delusional Disorder

**Fig. S1: Quality control pipeline for genetic analyses of the Mision Origen cohort.**
Flowchart summarizing the sequential quality control steps applied to exome sequencing data prior to association analyses. Sample counts at each step are shown in yellow rectangles; the number of variants retained at each step is shown in gray rectangles. After exome and sample QC, we performed QC on imputed data and restricted it to those samples that passed exome-based QC. Imputed genotype dataset was merged with the exome sequencing dataset, and for variants present in both datasets, the exome sequencing genotypes were preferentially retained.

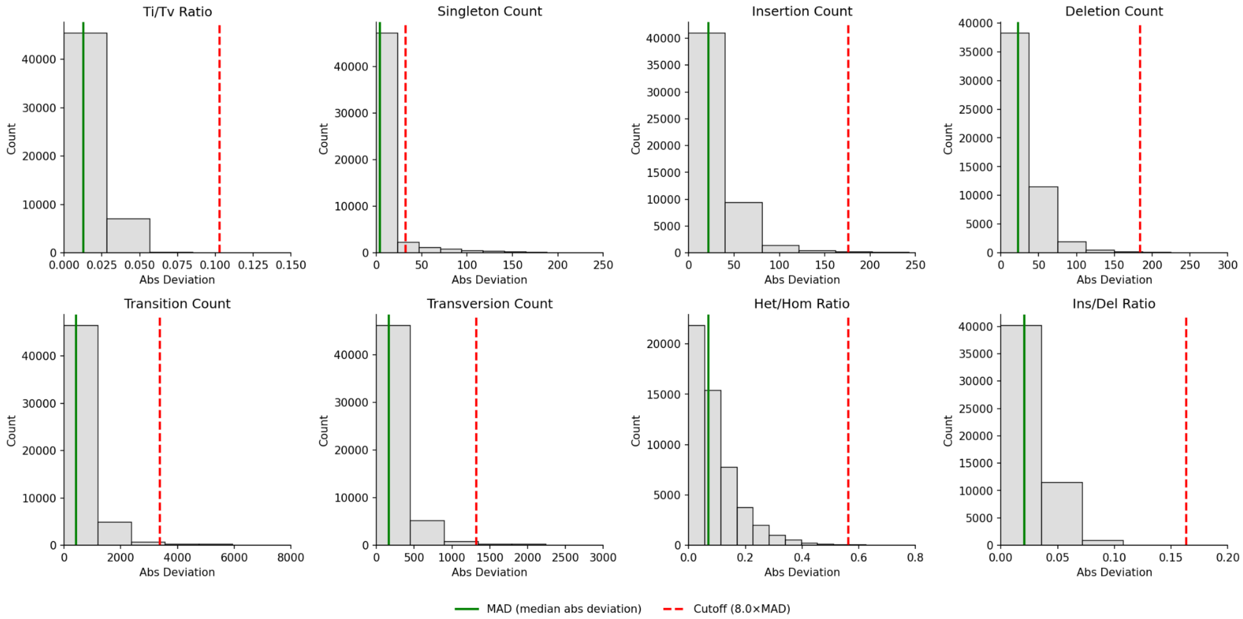

**Fig. S2**: **Sample quality control metrics from exome sequencing data.** Distribution of eight per-sample quality control metrics computed in Hail. For each metric, the median absolute deviation (MAD) is shown (green vertical line), with the exclusion threshold set at 8×MAD from the median (dashed red line). Samples failing all eight metrics simultaneously were excluded from downstream analyses.

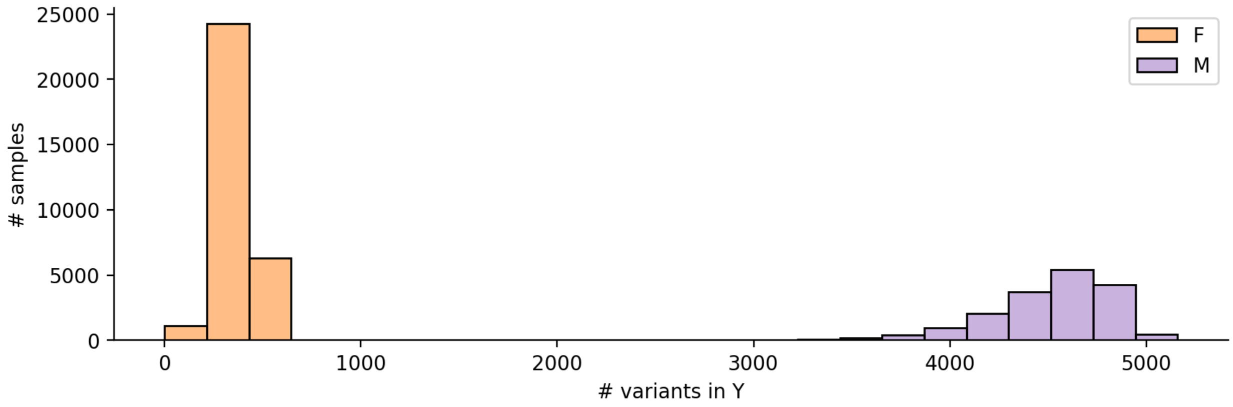

**Fig. S3: Distribution of variant counts on the Y chromosome used to assign genetic sex.** Per-sample count of variants on the Y chromosome, used to distinguish genetic males from genetic females. The bimodal distribution reflects the expected separation between individuals with (males) and without (females) a Y chromosome. Samples falling below the threshold of 1000 were assigned female, and samples above were assigned male.

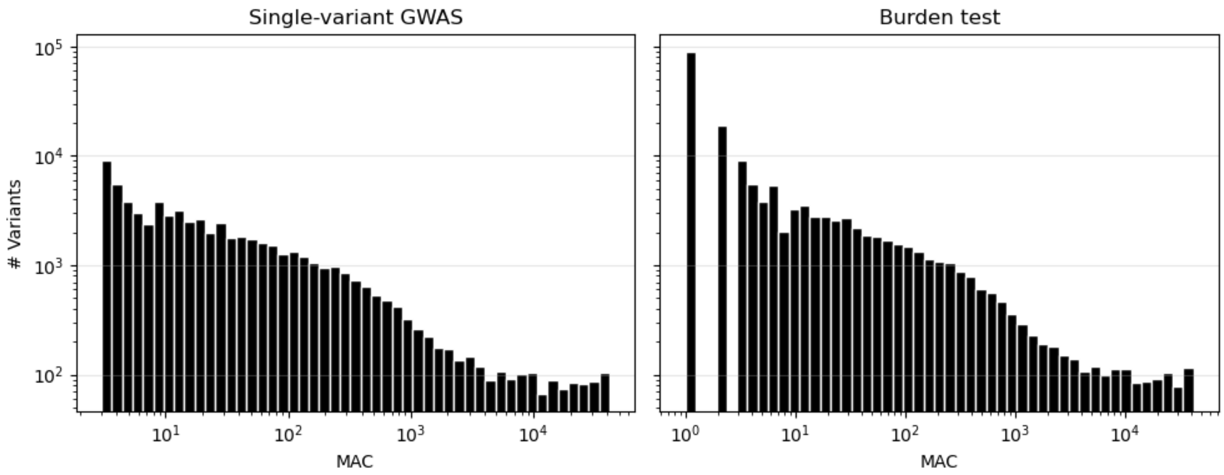

**Fig. S4: Distribution of minor allele count (MAC) of variants included in association analyses.** Per-variant minor allele count for variants included in the single-variant association test (left) and the gene-level burden test (right).

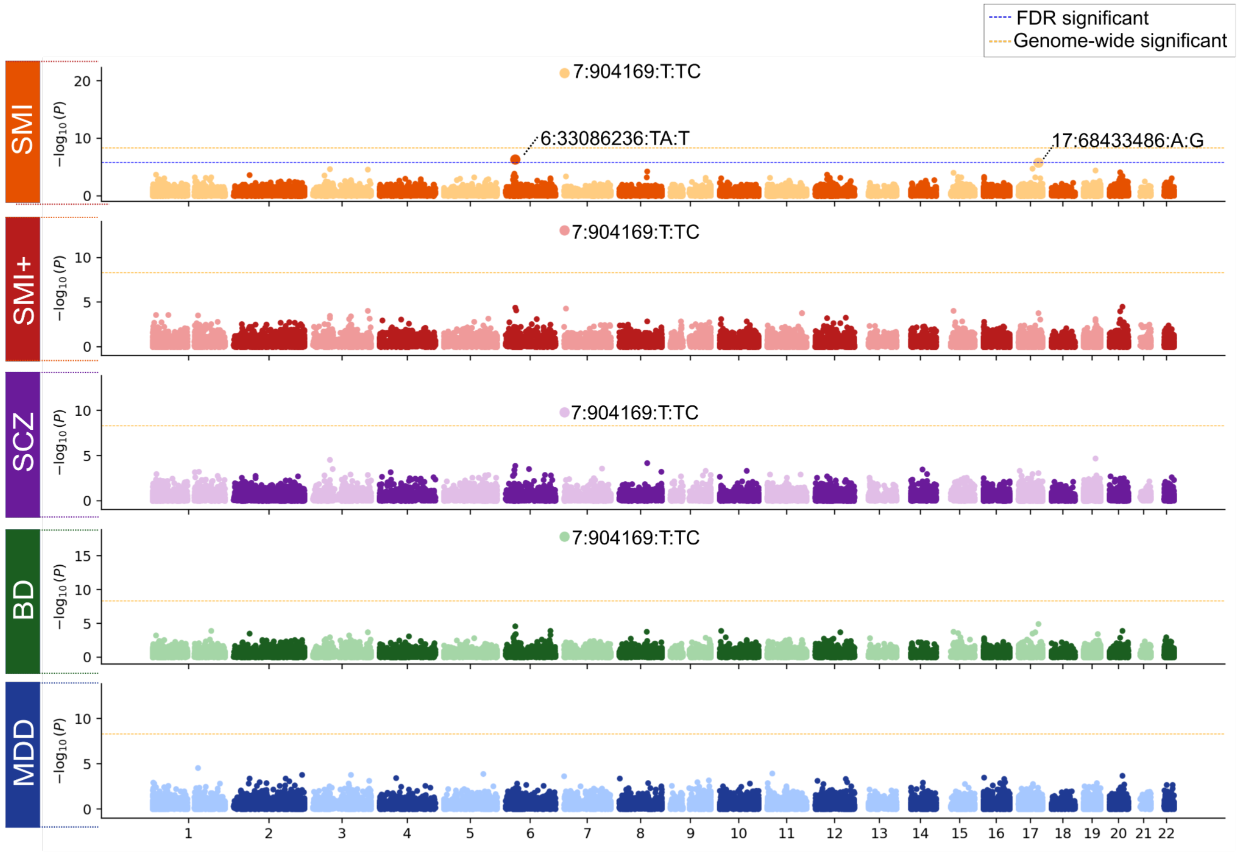

**Fig. S5: Deleterious single variant associations with SMI (and SMI+) phenotypes in Mision Origen.** Single-variant association of loss-of-function -and damaging missense variants analyzed using SAIGE logistic mixed models across five phenotypes: severe mental illness (SMI), SMI including major depressive disorder (SMI+), schizophrenia including schizoaffective disorder (SCZ), bipolar disorder (BD), and major depressive disorder (MDD). The FDR line is omitted for phenotypes in which only 7:904169:T:TC reached significance.

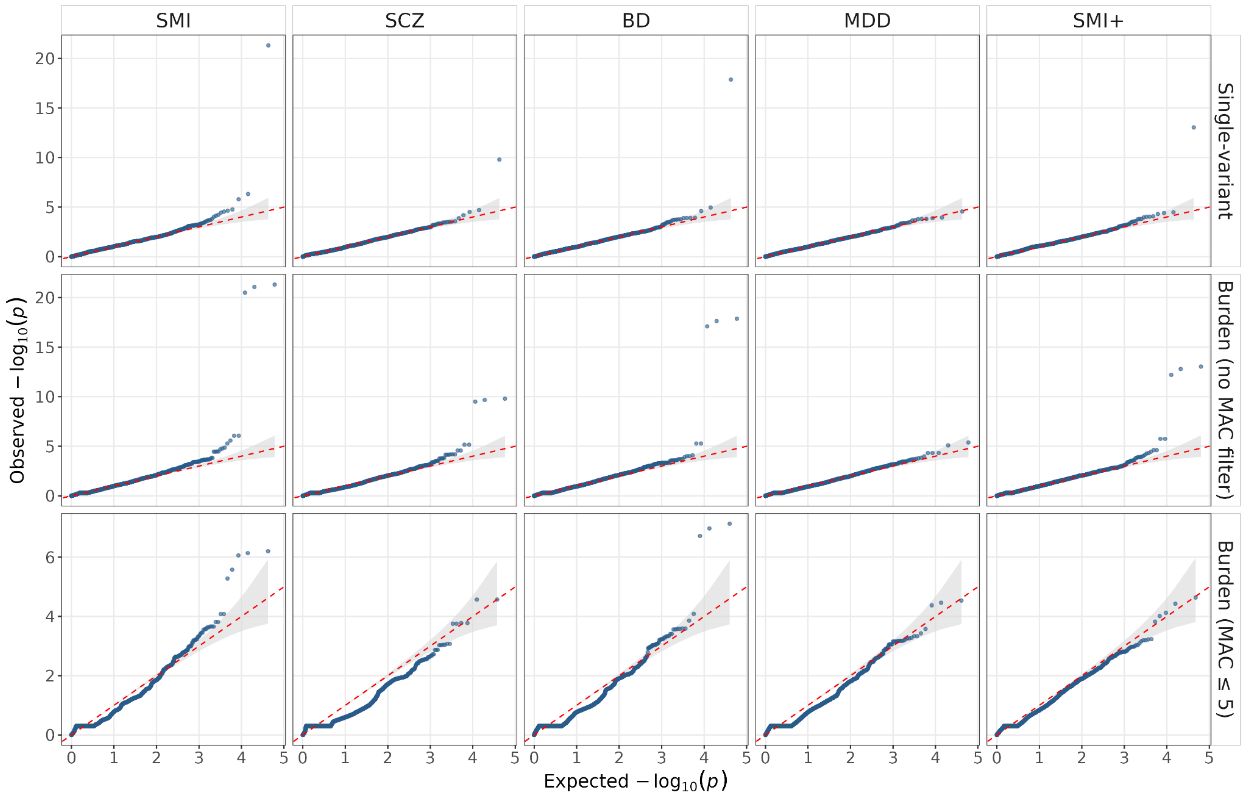

**Fig. S6: Q-Q plots for single-variant and gene-based (burden) associations.** Distributions of observed versus expected −log10(p-values) are shown for three analysis types (single-variant, burden [no MAC filter], and burden [MAC ≤ 5]) and five outcomes (SMI, SCZ, BD, MDD, and SMI+). The grey shaded region indicates the 95% confidence band under the null hypothesis of no association. The red dashed line indicates the expected distribution under the null (y = x).

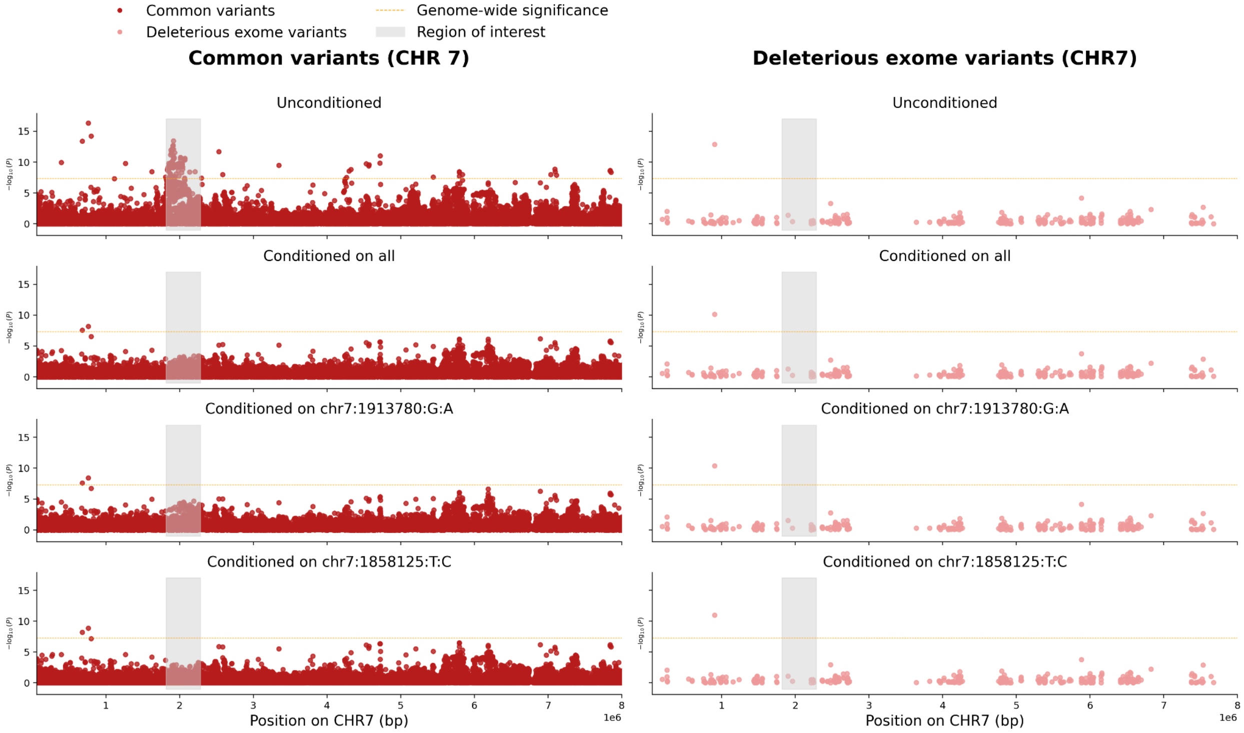

**Fig. S7: Conditional association analysis of variant 7:904169:T:TC on known chromosome 7 loci (1,817,033–2,283,308 basepairs) for SMI+.** Association results for common (left) and rare (right) variants restricted to chromosome 7 (positions 60,000–8,000,000). The top row shows unconditional associations; subsequent rows show results conditioned jointly and then individually on the two lead common-variant signals identified by LD clumping ($p < 5.00e\text{-}08$, $r^2 < 0.50$, ±500 kb window). The gray shaded area marks the region of interest identified in previous GWAS; the horizontal dashed line indicates common variant genome-wide significance threshold ($p < 5.00e\text{-}08$). Positions are on GRCh38; detailed results are in **Table S9**.

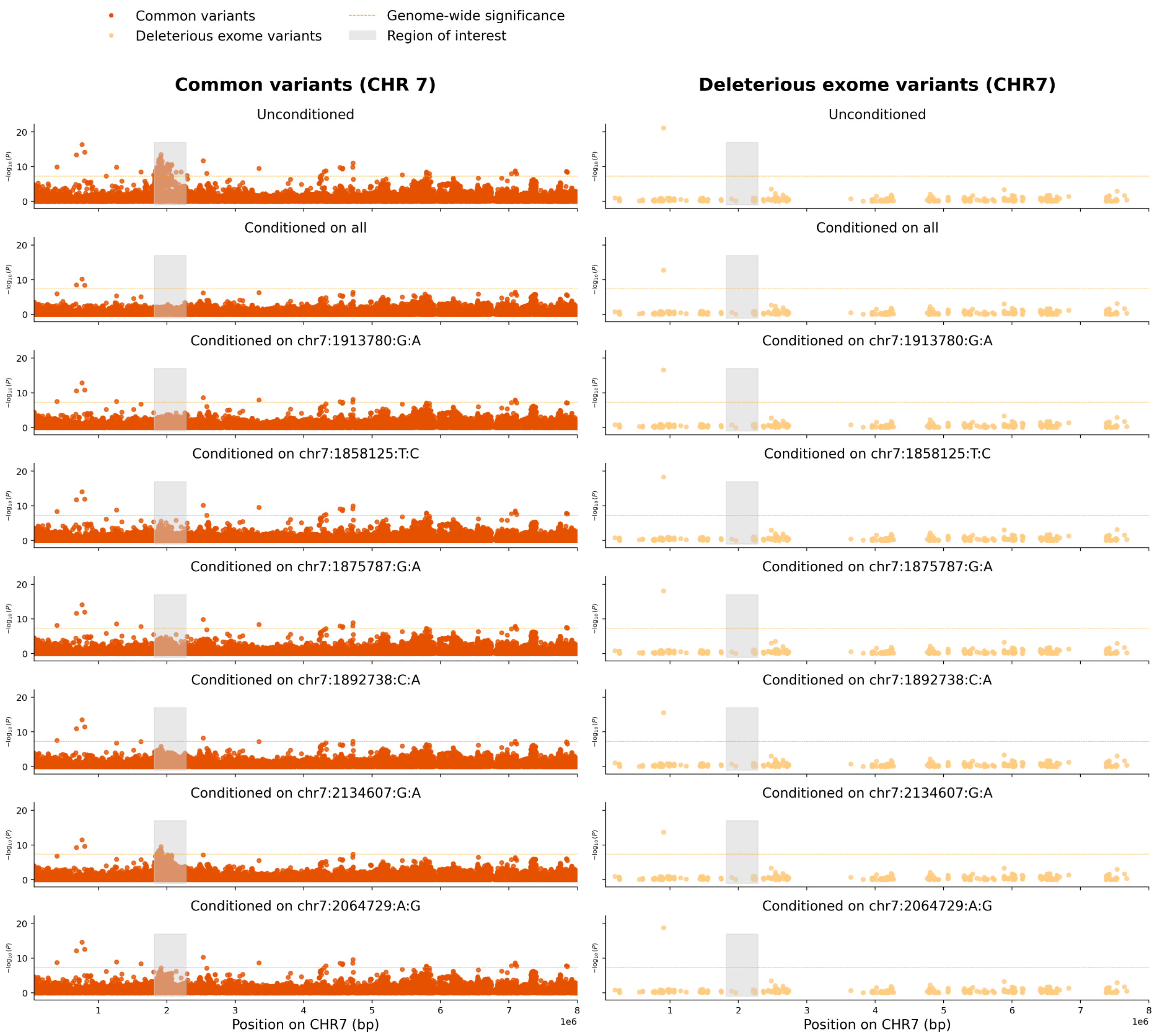

**Fig. S8: Conditional association analysis of variant 7:904169:T:TC on known chromosome 7 loci (1,817,033–2,283,308 basepairs) for SMI.** Association results for common (left) and rare (right) variants restricted to chromosome 7 (positions 60,000–8,000,000). The top row shows unconditional associations; subsequent rows show results conditioned jointly and then individually on the six lead common-variant signals identified by LD clumping ($p < 5.00e-08$, $r^2 < 0.50$, ±500 kb window). The gray shaded area marks the region of interest identified in previous GWAS[1–3,5]; the horizontal dashed line indicates common variant genome-wide significance threshold ($p < 5.00e-08$). Detailed results are available in **Table S9**.

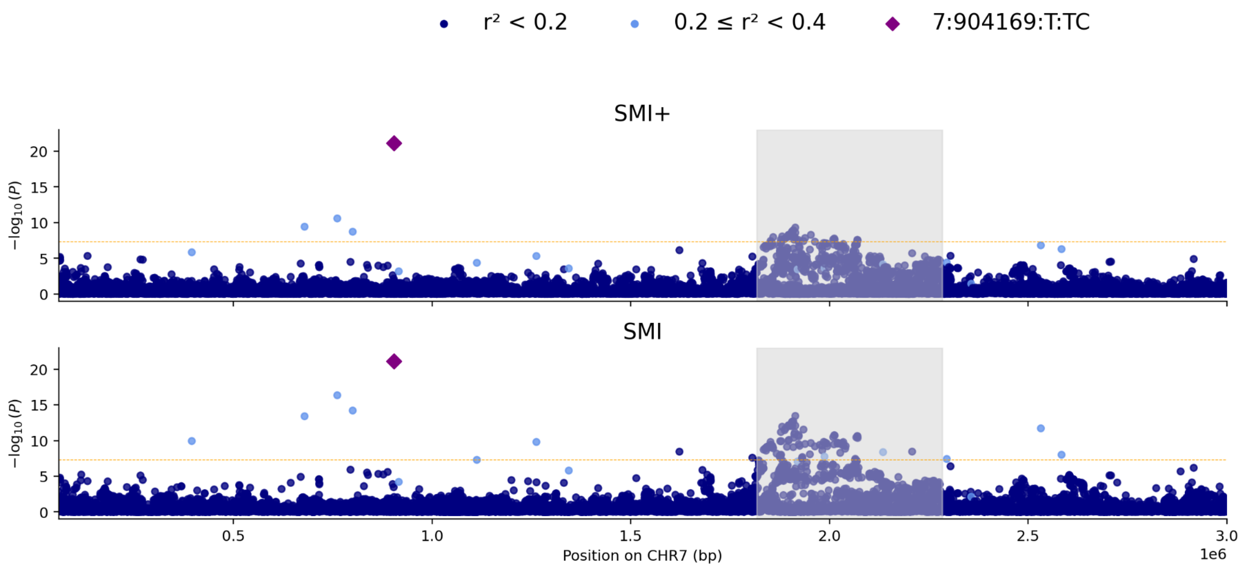

**Fig. S9: Common-variant association results in the ADAP1 locus, colored by linkage disequilibrium with 7:904169:T:TC variant.** Association results for common variants (MAF ≥ 0.01) on chromosome 7 (positions 60,000–6,000,000 basepairs) for severe mental illness (SMI+, top) and SMI (bottom). Each point represents a common variant coloured by $r^2$ with the 7:904169:T:TC variant, computed in the Mision Origen sample following the LocusZoom convention (navy: $r^2 < 0.2$; cornflower blue: 0.2–0.4; green: 0.4–0.6; orange: 0.6–0.8; red: ≥ 0.8). The 7:904169:T:TC variant is shown as a purple diamond. The shaded gray area indicates the region of interest with previously identified loci (7:1,817,033–2,283,308 basepairs)[1–3,5]. The horizontal dashed line indicates common variant genome-wide significance threshold ($p < 5.00e-08$). Results on genome-wide significant variants outside the region of interest are in Table S10. Positions are on GRCh38.

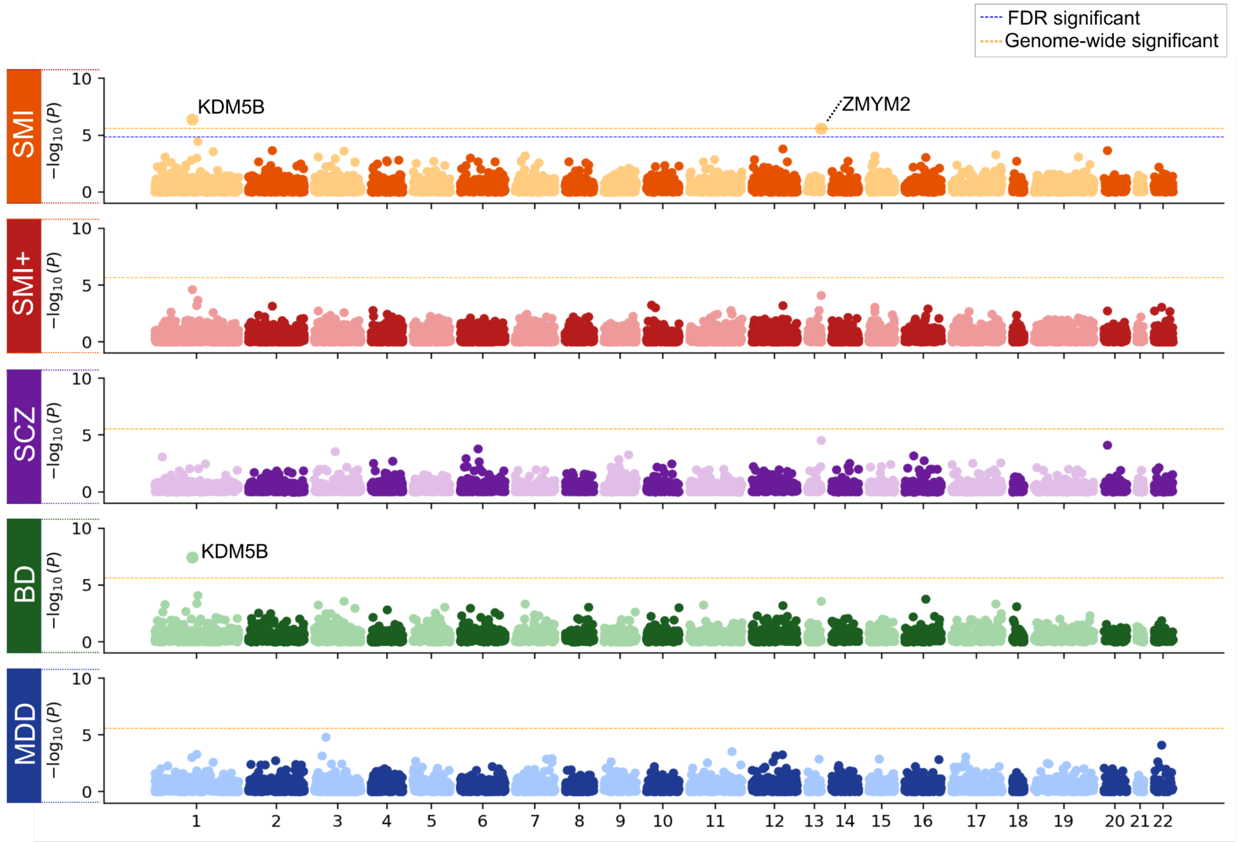

**Fig. S10: Burden test association results for damaging variants (MAC ≤ 5).** Gene-level burden test results for protein-truncating and damaging missense variants with minor allele count ≤ 5, tested across five phenotypes (SMI, SMI+, SCZ, BD, and MDD). Each point represents a gene, plotted by chromosomal position against $-\log_{10}$(p-value). FDR lines are omitted where the FDR threshold exceeded the exome-wide threshold on the -log10 scale.

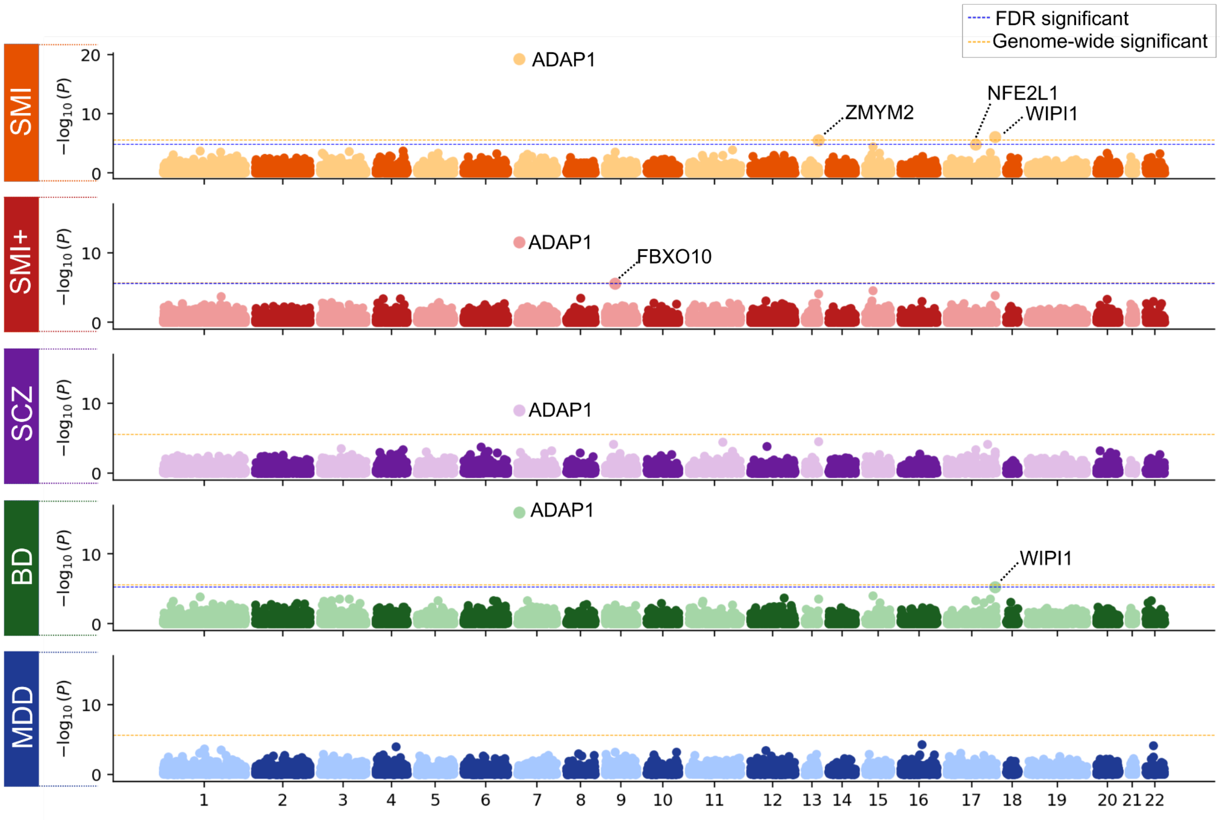

**Fig. S11: Burden test association results for damaging variants (no MAC filter).** Gene-level burden test results for protein-truncating and damaging missense variants, tested across five phenotypes (SMI, SMI+, SCZ, BD, and MDD). Each point represents a gene, plotted by chromosomal position against $-\log_{10}$(p-value). FDR significance lines are omitted where the FDR threshold exceeded the genome-wide threshold on the -log10 scale.

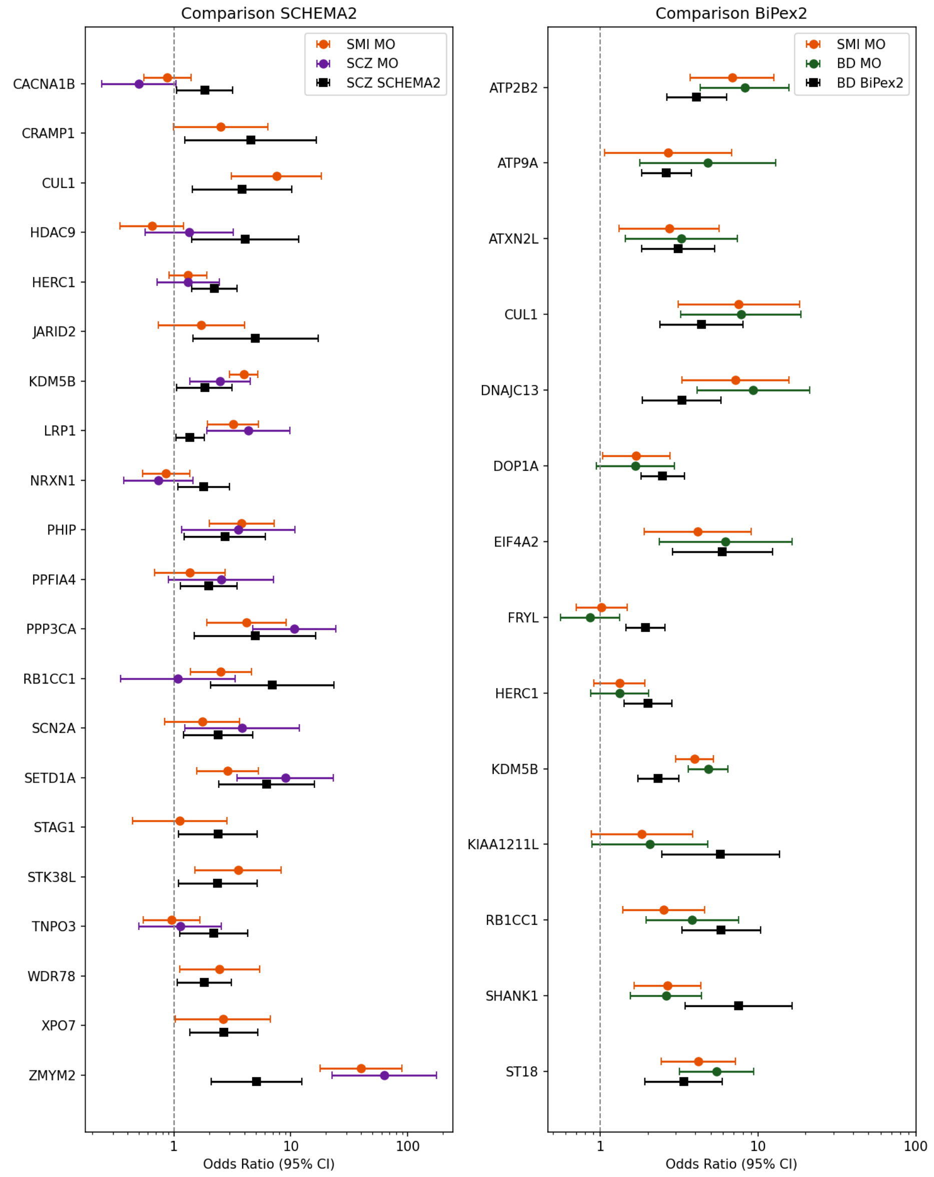

**Fig. S12: Odds ratio comparison with SCHEMA2 and BipEx2 consortia results for damaging variants with a minor allele count (MAC) ≤ 5.** Genes shown are those reaching genome-wide or FDR significance in SCHEMA2 or BiPEx2 that had deleterious variants with MAC<=5 in our sample. Genes reported in SCHEMA2 or BiPex2 but without qualifying variants in our dataset, and therefore not shown, were: *AKAP11, ATP1A1, CHRM4, DAGLA, FYN, LRRC4, RFX3, SCAF1, SP4, UBE2E3, ZC3H12B, ZMYND11* for SCHEMA2 and *AKAP11, AKR7L, C1orf61, EIF4E2, HDAC3, HECTD2, HEPACAM, HES4, KLF1, NBPF14, PCDHGA8, RAB3D, SLC2A11, SP4, SYT1, TCF7L1, TERF2, TEX261, TOPAZ1* for BiPex2. Note that there is substantial sample overlap between Mision Origen (MO) and both consortia: Mision Origen contributed n=19,424 participants to SCHEMA2 (8% of the total sample) and n=27,378 participants to BipEx2 (12% of the total sample).

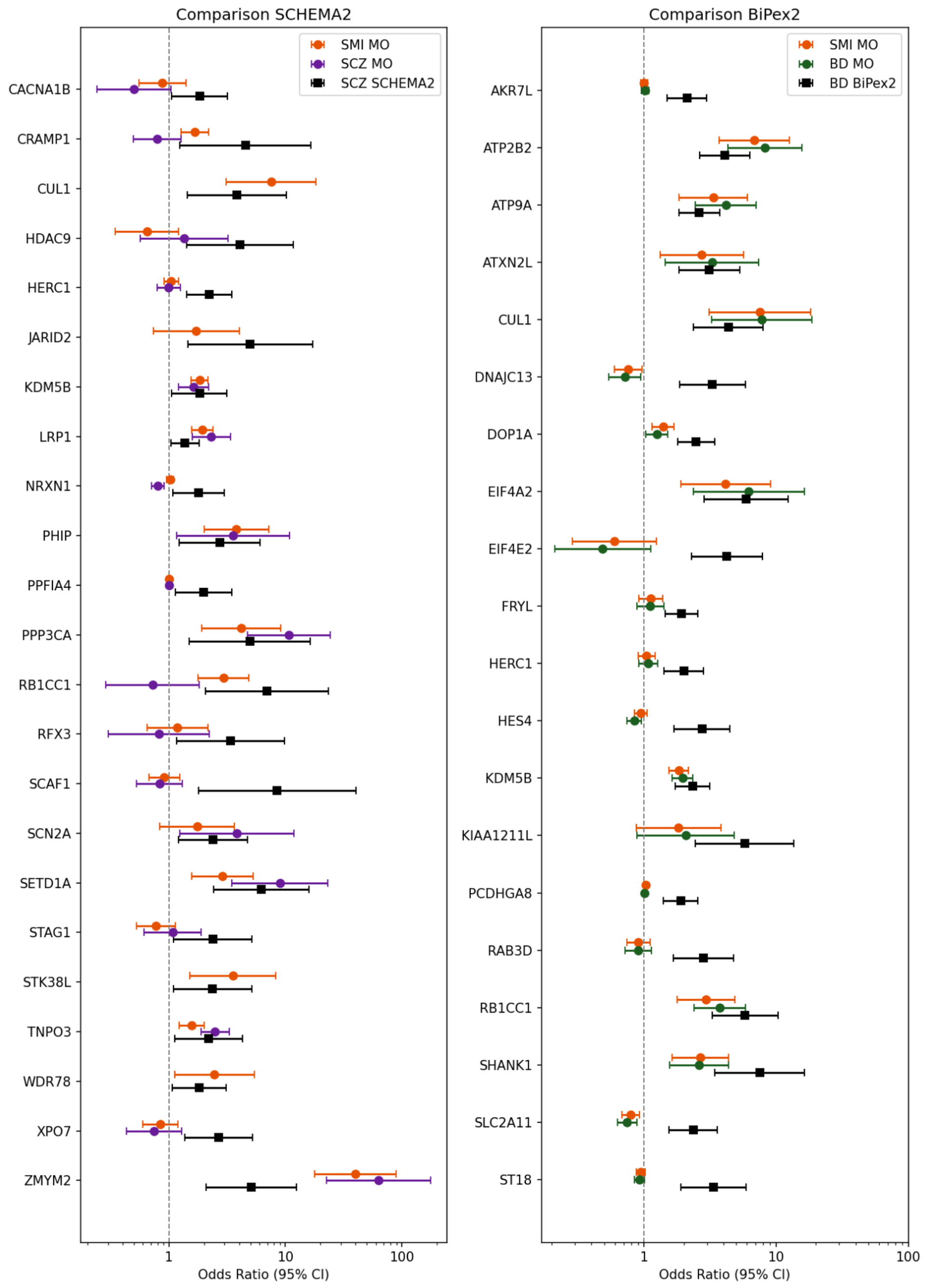

**Fig. S13: Odds ratio comparison with SCHEMA2 and BipEx2 consortia results for damaging variants without any MAC filter.** Genes shown are those reaching genome-wide or FDR significance in SCHEMA2 or BiPEx2 that had deleterious variants. Genes reported in SCHEMA2 or BiPex2 but without qualifying variants in our dataset, and therefore not shown, were: *AKAP11, ATP1A1, CHRM4, DAGLA, FYN, LRRC4, SP4, UBE2E3, ZC3H12B, ZMYND11* for SCHEMA2 and *AKAP11, C1orf61, HDAC3, HECTD2, HEPACAM, KLF1, NBPF14*, *SP4, SYT1, TCF7L1, TERF2, TEX261, TOPAZ1* for BiPex2. Note that there is substantial sample overlap between Mision Origen (MO) and both consortia: Mision Origen contributed n=19,424 participants to SCHEMA2 (8% of the total sample) and n=27,378 participants to BipEx2 (12% of the total sample).

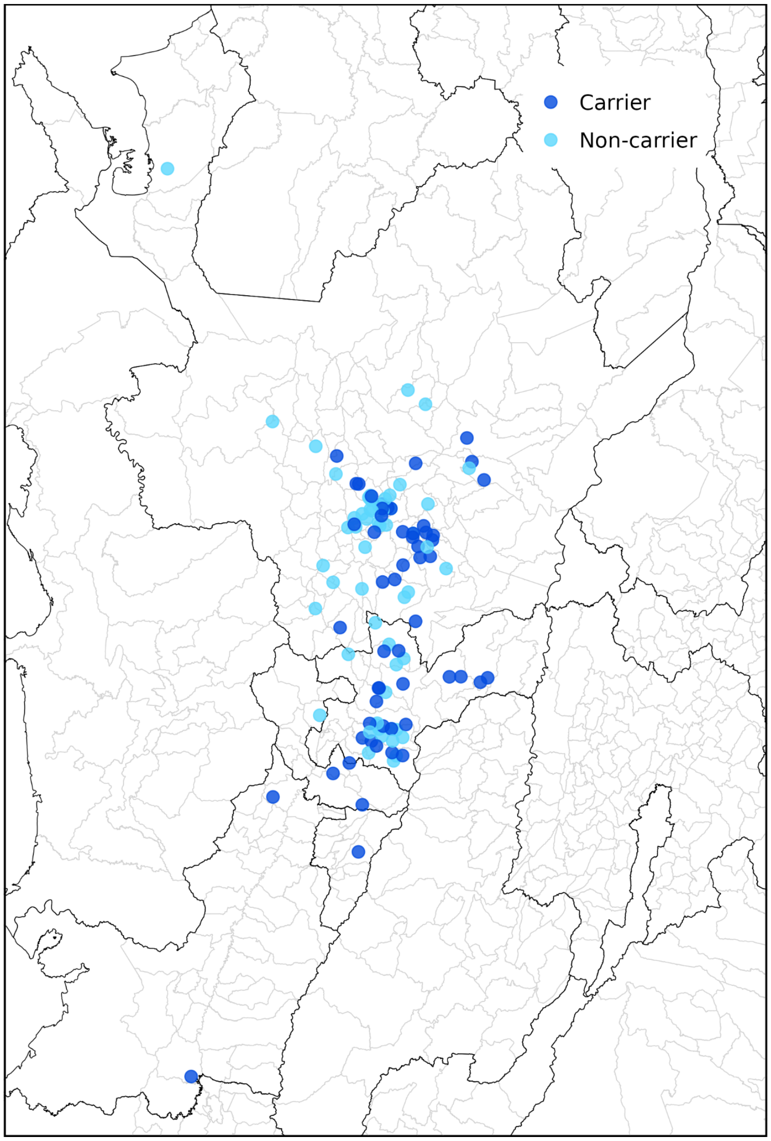

**Fig. S14: Geographic distribution of 17:68433486:A:G variant carriers.** Distribution of 17:68433486:A:G variant carriers (n=56) and equal number of randomly selected non-carriers. To protect participant confidentiality, locations are jittered around city centers and do not reflect addresses.

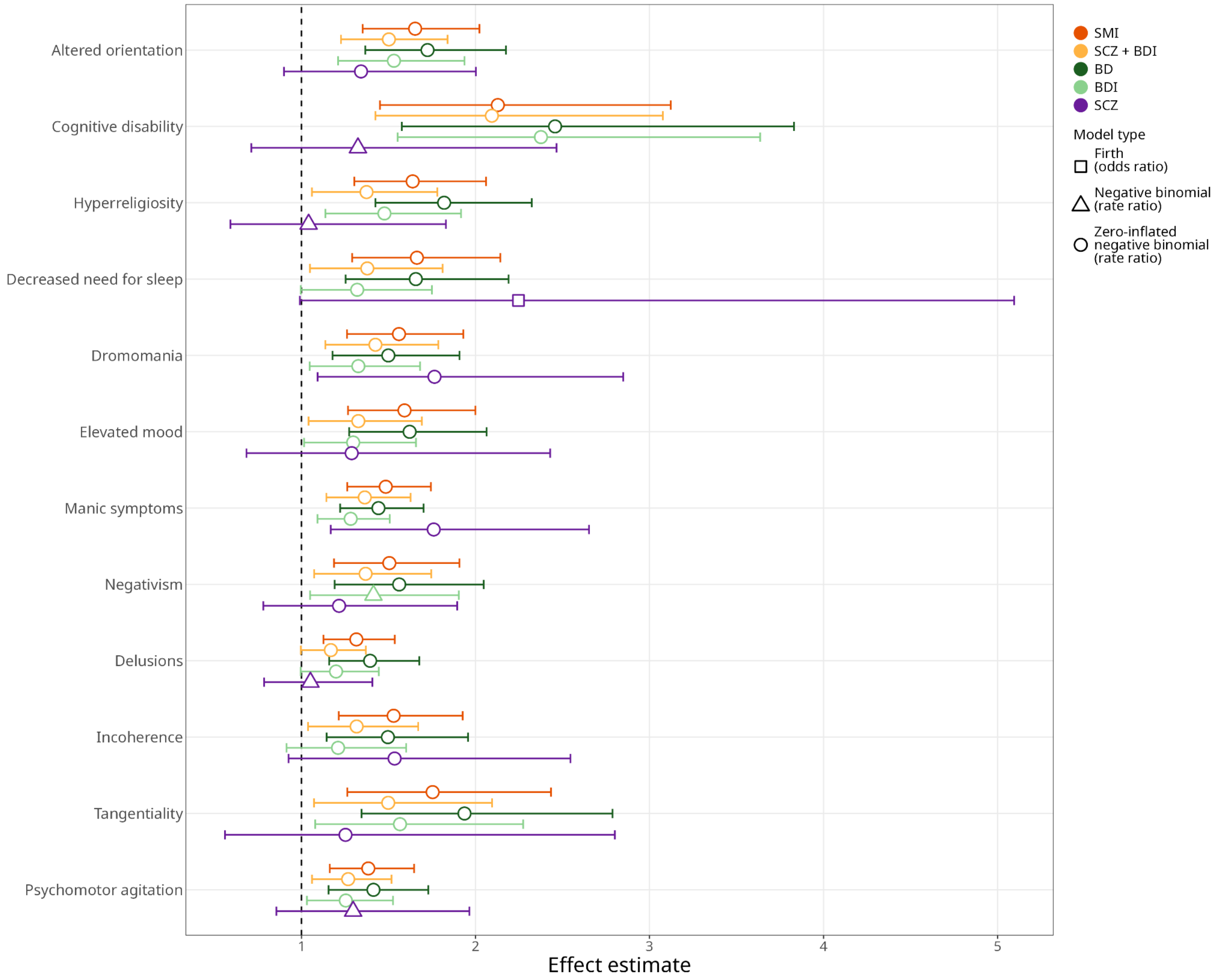

**Fig. S15: Sensitivity analysis: effect of 7:904169:T:TC carrier status on item-level phenotypes in participants with the following diagnoses: SMI, SCZ + BD1, BD, BD1, and SCZ.** Univariate analyses were conducted within each diagnosis subset for phenotypes that were significant in the main phenotypic analysis (highlighted in dark orange). Effect estimates are shown with non-carriers as the reference group. Item-level phenotypes were modeled as rates using negative binomial or zero-inflated negative binomial regression, selected by likelihood ratio test; sparse phenotypes (<15% of participants endorse the phenotype) were modeled as binary outcomes using Firth logistic regression. Rate ratios and odds ratios are shown with bootstrap 95% confidence intervals.

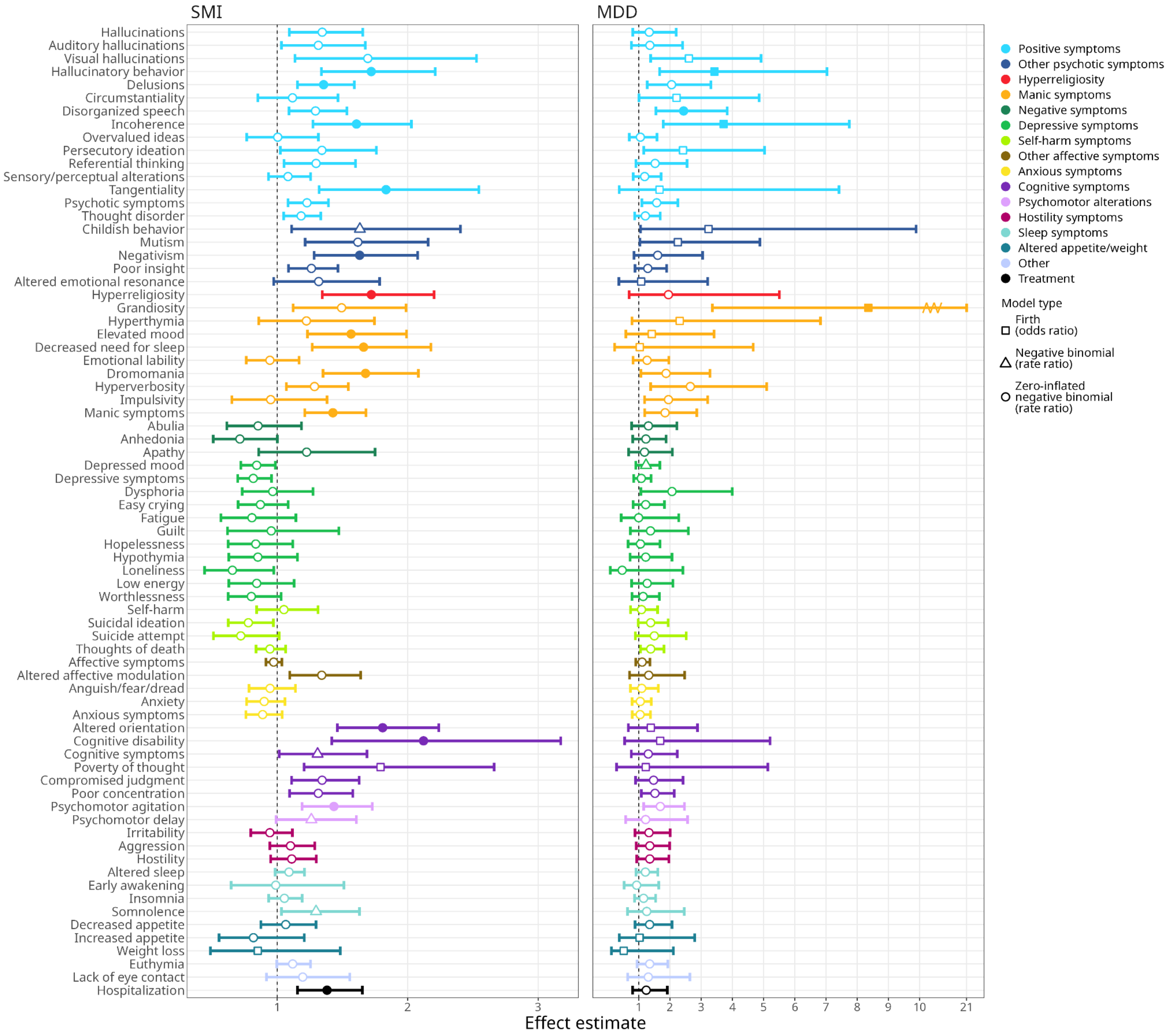

**Fig. S16: Sensitivity analysis: item-level phenotypic analyses, adjusting for the first five genetic principal components.** Effect estimates from univariate associations between 7:904169:T:TC carrier status and item-level phenotypes shown for participants with SMI (left) and participants with MDD (right), with non-carriers as the reference group. Item-level phenotypes were modeled as rates using negative binomial or zero-inflated negative binomial regression, selected by likelihood ratio test; sparse phenotypes (<15% of participants endorsing the phenotype) were modeled as binary outcomes using Firth logistic regression. Rate ratios and odds ratios are shown with bootstrap 95% confidence intervals. Filled points indicate associations surviving Bonferroni correction.

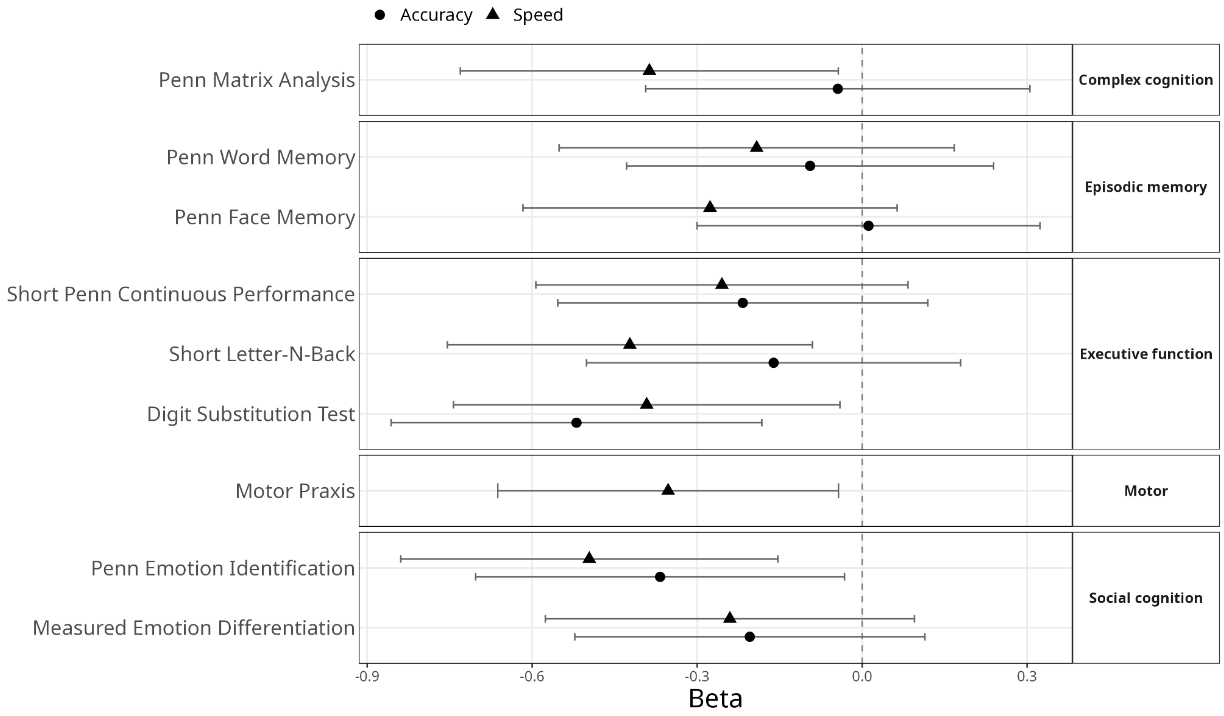

**Fig. S17: Effect of carrier status on cognitive function in participants with SMI diagnoses**. Speed and accuracy of cognition in five domains was modeled as a function of 7:904169:T:TC carrier status, with non-carrier as reference. Beta estimates cognition of carriers, in standard deviation units; with negative values indicating reduced cognitive function in carriers compared to non-carriers. Error bars are the 95% CI. The dashed line at zero represents the null hypothesis of cognition in carriers equal to cognition in non-carriers. Prior to analysis, cognition had been regressed on age, education, and sex, and residuals were quantile-normal transformed. Confidence intervals that do not overlap zero are significant at the 0.05 level, one test (accuracy of the Digit Substitution test) remains significant when controlling for multiple testing using a Bonferroni correction for 17 tests. The number of carriers for each test are shown in Table S26.

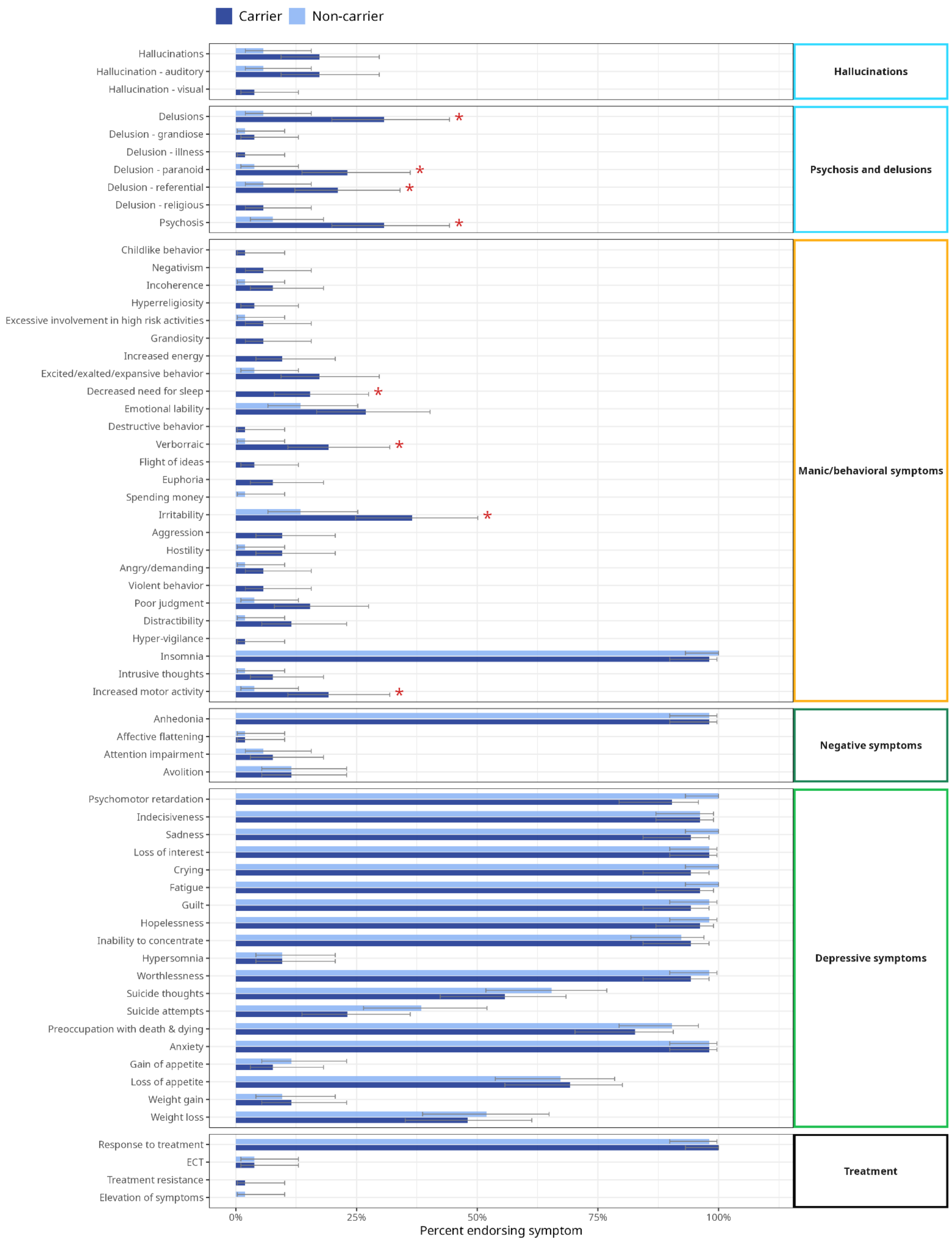

**Fig. S18: Clinical feature endorsement in paired MDD carriers and non-carriers from structured chart review.** Proportion of participants endorsing each clinical feature, derived from structured chart review, compared between MDD carriers and paired non-carriers, shown with Wilson 95% confidence intervals. Nominal significance from Chi-squared test denoted with a red asterisk. Features endorsed by 0% or 100% of both carriers and non-carriers were excluded from the plot.

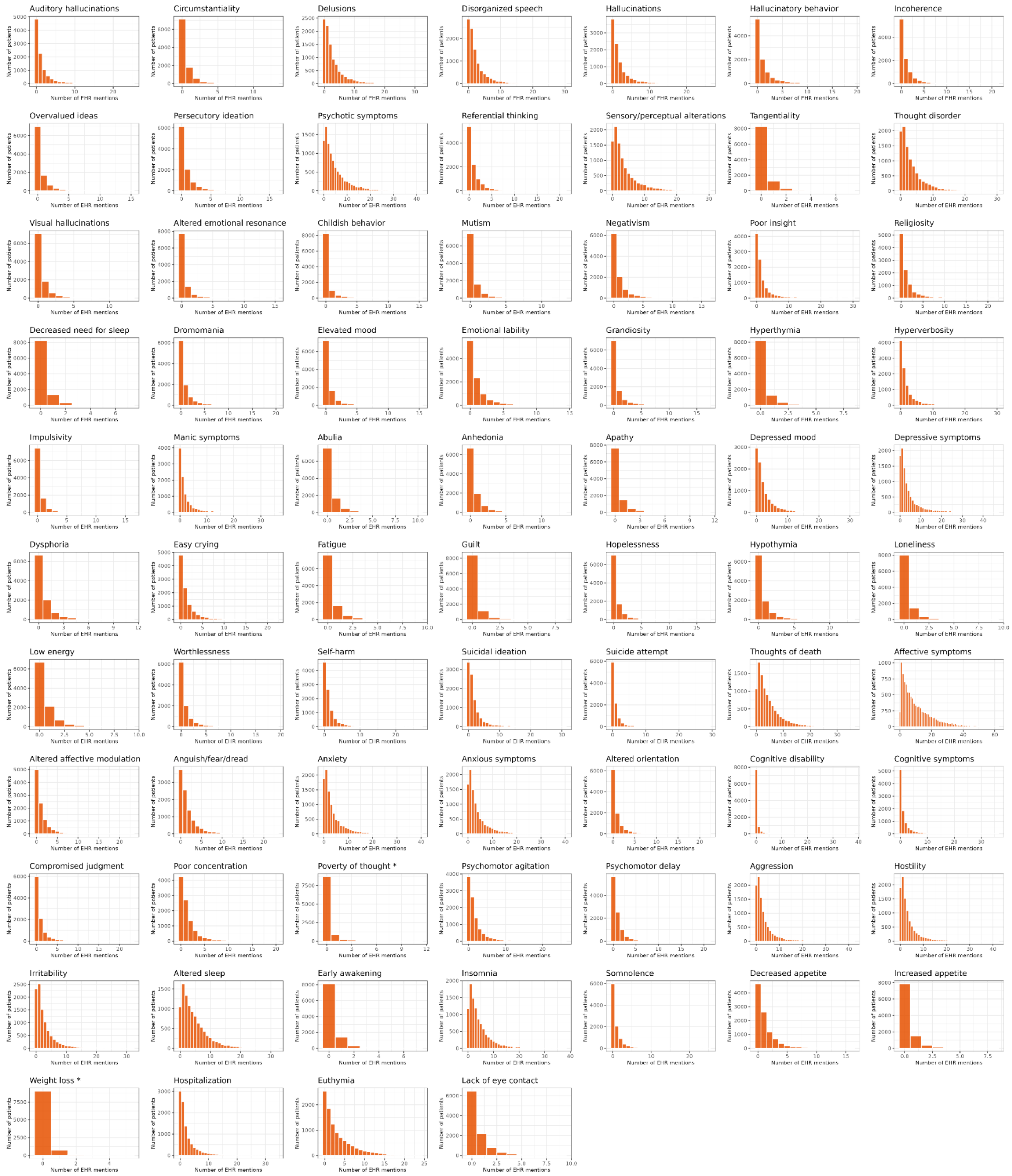

**Fig. S19: Item-level phenotype count distributions for participants with SMI.** Distributions of phenotype counts (occurrences in the EHR) for participants with SMI. Phenotypes models as binary features shown with asterisks.

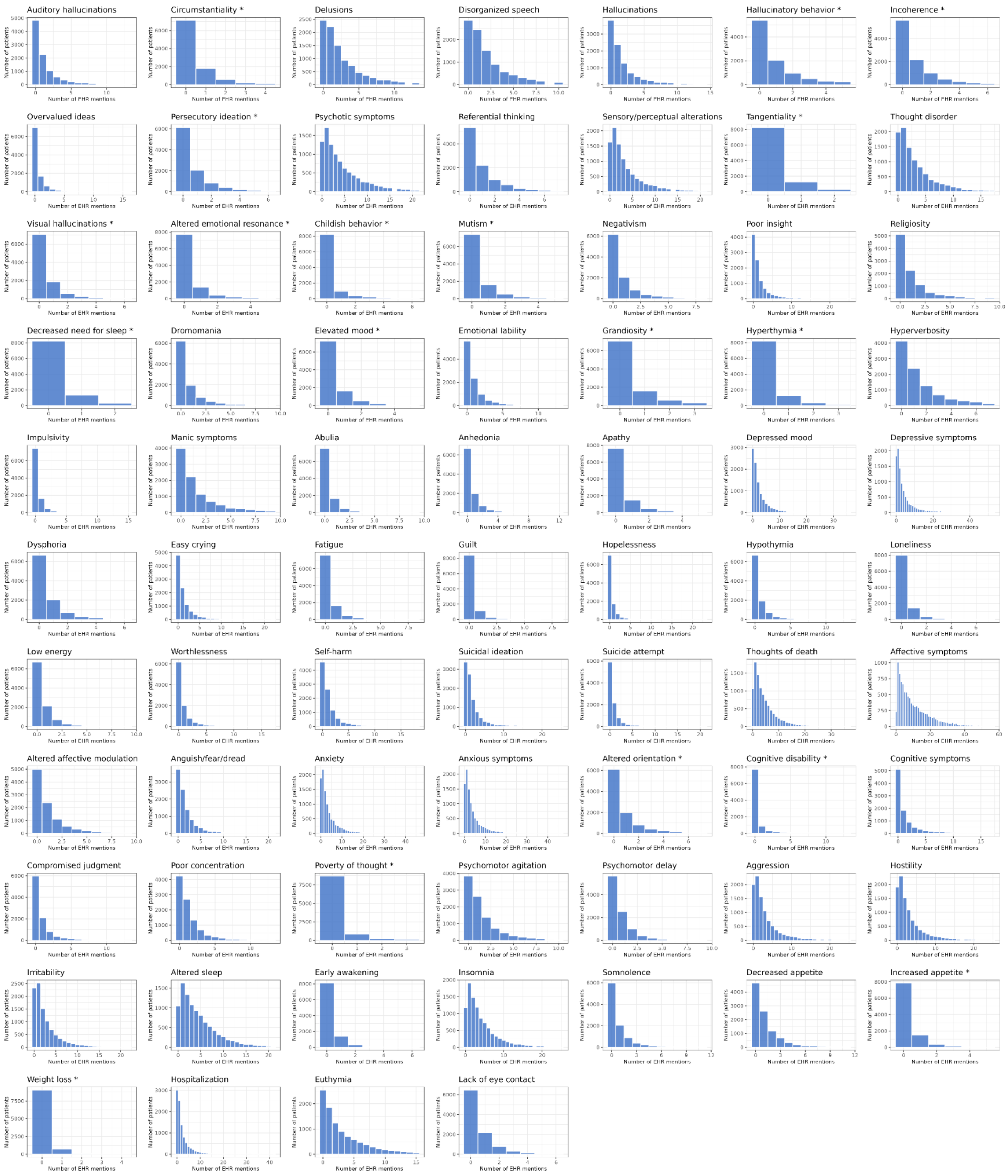

**Fig. S20: Item-level phenotype count distributions for participants with MDD.** Distributions of phenotype counts (occurrences in the EHR) for participants with MDD. Phenotypes models as binary features shown with asterisks.

#### Supplementary Table Information

**Table S1. ICD-10 codes used for SMI attribution in Mision Origen**

**Table S2. Number of study participants, stratified by diagnosis and recruitment site**

**Table S3. Deleterious variants association results for SMI.** Results are reported for loss-of-function and damaging missense variants (MAC ≥ 3) tested for association with severe mental illness (SMI). Associations were tested using SAIGE logistic mixed models with a sparse genetic relatedness matrix to account for population structure and relatedness, with the first five principal components of ancestry as covariates. Genome-wide significance was defined as p < 5.00e-09, and FDR significance was defined at a 5% false discovery rate using the Benjamini-Hochberg procedure p ≤ 1.62e-06.

**Table S4. Deleterious variants association results for SMI+.** Results are reported for protein-truncating and damaging missense variants (MAC ≥ 3) tested for association with severe mental illness (SMI+). Associations were tested using SAIGE logistic mixed models with a sparse genetic relatedness matrix to account for population structure and relatedness, with the first five principal components of ancestry as covariates. Genome-wide significance was defined as p < 5.00e-09, and FDR significance was defined at a 5% false discovery rate using the Benjamini-Hochberg procedure as p ≤ 9.16e-14.

**Table S5. Deleterious variants association results for SCZ.** Results are reported for protein-truncating and damaging missense variants (MAC ≥ 3) tested for association with severe mental illness (SCZ). Associations were tested using SAIGE logistic mixed models with a sparse genetic relatedness matrix to account for population structure and relatedness, with the first five principal components of ancestry as covariates. Genome-wide significance was defined as p < 5.00e-09, and FDR significance was defined at a 5% false discovery rate using the Benjamini-Hochberg procedure as p ≤ 1.61e-10.

**Table S6. Deleterious variants association results for BD.** Results are reported for protein-truncating and damaging missense variants (MAC ≥ 3) tested for association with severe mental illness (BD). Associations were tested using SAIGE logistic mixed models with a sparse genetic relatedness matrix to account for population structure and relatedness, with the first five principal components of ancestry as covariates. Genome-wide significance was defined as p < 5.00e-09, and FDR significance was defined at a 5% false discovery rate using the Benjamini-Hochberg procedure as p ≤ 1.36e-18.

**Table S7. Deleterious variants association results for MDD.** Results are reported for protein-truncating and damaging missense variants (MAC ≥ 3) tested for association with severe mental illness (MDD). Associations were tested using SAIGE logistic mixed models with a sparse genetic relatedness matrix to account for population structure and relatedness, with the first five principal components of ancestry as covariates. Genome-wide significance was defined as p < 5.00e-09, and FDR significance was defined at a 5% false discovery rate using the Benjamini-Hochberg procedure. No variants reached FDR significance for MDD.

**Table S8. Number of carriers of genome-wide and FDR- significant deleterious variants in single-variant associations with diagnosis, stratified by zygosity.** Het = heterozygous, Hom = homozygous.

**Table S9. The 7:904169:T:TC variant remains genome-wide significant after conditioning on known SMI-associated loci.** (a) Significant independent loci within the chromosome 7 region previously associated with SMI and SMI+ (7:1,817,033–2,283,308) using LD clumping ($p < 5.00e-08$, $r^2 < 0.50$, ±500 kb window). (b) Results for 7:904169:T:TC variant after conditioning loci from Table (a) independently and together (all).

**Table S10.The 7:904169:T:TC variant remains genome-wide significant after conditioning on newly identified associations to common variants** (a) Common variants (MAF ≥ 1%) in chromosome 7 that are genome-wide significant in association with SMI+ or SMI are in weak LD with 7:904169:T:TC variant. (b) Results for 7:904169:T:TC variant after conditioning loci from Table a.

**Table S11. Deleterious variants burden test results in association with SMI, including all variants regardless of MAC.** Results are reported for protein-truncating and damaging missense variants tested for association with severe mental illness (SMI). Associations were tested using SAIGE-GENE logistic mixed models burden test with a sparse genetic relatedness matrix to account for population structure and relatedness, with the first five principal components of ancestry as covariates. Genome-wide significance was defined using Bonferroni correction at an alpha level of 0.05, considering all genes tested across both variant filters (no filter and MAC ≤ 5): $p \leq 2.51e-06$. FDR significance was similarly defined at the phenotype level, pooling p-values across both variant filters and applying a 5% false discovery rate using the Benjamini-Hochberg procedure: $p \leq 1.38e-05$.

**Table S12. Deleterious variants burden test results in association with SMI+, including all variants regardless of MAC.** Results are reported for protein-truncating and damaging missense variants tested for association with severe mental illness including Major Depressive Disorder (SMI+). Associations were tested using SAIGE-GENE logistic mixed models burden test with a sparse genetic relatedness matrix to account for population structure and relatedness, with the first five principal components of ancestry as covariates. Genome-wide significance was defined using Bonferroni correction at an alpha level of 0.05, considering all genes tested across both variant filters (no filter and MAC ≤ 5): $p \leq 2.30e-06$. FDR significance was similarly defined at the phenotype level, pooling p-values across both variant filters and applying a 5% false discovery rate using the Benjamini-Hochberg procedure: $p \leq 2.83e-06$.

**Table S13. Deleterious variants burden test results in association with SCZ, including all variants regardless of MAC.** Results are reported for protein-truncating and damaging missense variants tested for association with schizophrenia (SCZ). Associations were tested using SAIGE-GENE logistic mixed models burden test with a sparse genetic relatedness matrix to account for population structure and relatedness, with the first five principal components of ancestry as covariates. Genome-wide significance was defined using Bonferroni correction at an alpha level of 0.05, considering all genes tested across both variant filters (no filter and MAC ≤ 5): $p \leq 2.80e-06$. FDR significance was similarly defined at the phenotype level, pooling p-values across both variant filters and applying a 5% false discovery rate using the Benjamini-Hochberg procedure: $p \leq 1.05e-09$.

**Table S14. Deleterious variants burden test results in association with BD, including all variants regardless of MAC.** Results are reported for protein-truncating and damaging missense variants tested for association with bipolar disorder (BD). Associations were tested using SAIGE-GENE logistic mixed models burden test with a sparse genetic relatedness matrix to account for population structure and relatedness, with the first five principal components of ancestry as covariates. Genome-wide significance was defined using Bonferroni correction at an alpha level of 0.05, considering all genes tested across both variant filters (no filter and MAC ≤ 5): $p \leq 2.63e-06$. FDR significance was similarly defined at the phenotype level, pooling p-values across both variant filters and applying a 5% false discovery rate using the Benjamini-Hochberg procedure: $p \leq 5.28e-06$.

**Table S15. Deleterious variants burden test results in association with MDD, including all variants regardless of MAC.** Results are reported for protein-truncating and damaging missense variants tested for association with Major Depressive Disorder (MDD). Associations were tested using SAIGE-GENE logistic mixed models burden test with a sparse genetic relatedness matrix to account for population structure and relatedness, with the first five principal components of ancestry as covariates. Genome-wide significance was defined using Bonferroni correction at an alpha level of 0.05, considering all genes tested across both variant filters (no filter and MAC ≤ 5): $p \leq 2.57e-06$. FDR significance was similarly defined at the phenotype level, pooling p-values across both variant filters and applying a 5% false discovery rate using the Benjamini-Hochberg procedure. No gene reached FDR significance for MDD.

**Table S16. Deleterious variants burden test results in association with SMI, including all variants with MAC≤5. Results are reported for protein-truncating and damaging missense variants tested for association with severe mental illness (SMI).** Associations were tested using SAIGE-GENE logistic mixed models burden test with a sparse genetic relatedness matrix to account for population structure and relatedness, with the first five principal components of ancestry as covariates. Genome-wide significance was defined using Bonferroni correction at an alpha level of 0.05, considering all genes tested across both variant filters (no filter and MAC ≤ 5): $p \leq 2.51e-06$. FDR significance was similarly defined at the phenotype level, pooling p-values across both variant filters and applying a 5% false discovery rate using the Benjamini-Hochberg procedure: $p \leq 1.38e-05$.

**Table S17. Deleterious variants burden test results in association with SMI+, including all variants with MAC ≤ 5.** Results are reported for protein-truncating and damaging missense variants tested for association with severe mental illness including Major Depressive Disorder (SMI+). Associations were tested using SAIGE-GENE logistic mixed models burden test with a sparse genetic relatedness matrix to account for population structure and relatedness, with the first five principal components of ancestry as covariates. Genome-wide significance was defined using Bonferroni correction at an alpha level of 0.05, considering all genes tested across both variant filters (no filter and MAC ≤ 5): $p \leq 2.30e-06$. FDR significance was similarly defined at the phenotype level, pooling p-values across both variant filters and applying a 5% false discovery rate using the Benjamini-Hochberg procedure:$p \leq 2.83e-06$.

**Table S18. Deleterious variants burden test results in association with SCZ, including all variants with MAC ≤ 5.** Results are reported for protein-truncating and damaging missense variants tested for association with schizophrenia (SCZ). Associations were tested using SAIGE-GENE logistic mixed models burden test with a sparse genetic relatedness matrix to account for population structure and relatedness, with the first five principal components of ancestry as covariates. Genome-wide significance was defined using Bonferroni correction at an alpha level of 0.05, considering all genes tested across both variant filters (no filter and MAC ≤ 5):  p ≤ 2.80e-06. FDR significance was similarly defined at the phenotype level, pooling p-values across both variant filters and applying a 5% false discovery rate using the Benjamini-Hochberg procedure: p ≤ 1.05e-09.

**Table S19. Deleterious variants burden test results in association with BD, including all variants with MAC ≤ 5.** Results are reported for protein-truncating and damaging missense variants tested for association with bipolar disorder (BD). Associations were tested using SAIGE-GENE logistic mixed models burden test with a sparse genetic relatedness matrix to account for population structure and relatedness, with the first five principal components of ancestry as covariates. Genome-wide significance was defined using Bonferroni correction at an alpha level of 0.05, considering all genes tested across both variant filters (no filter and MAC ≤ 5):  p ≤ 2.63e-06. FDR significance was similarly defined at the phenotype level, pooling p-values across both variant filters and applying a 5% false discovery rate using the Benjamini-Hochberg procedure: p ≤ 5.28e-06.

**Table S20. Deleterious variants burden test results in association with MDD, including all variants with MAC ≤ 5.** Results are reported for protein-truncating and damaging missense variants tested for association with Major Depressive Disorder (MDD). Associations were tested using SAIGE-GENE logistic mixed models burden test with a sparse genetic relatedness matrix to account for population structure and relatedness, with the first five principal components of ancestry as covariates. Genome-wide significance was defined using Bonferroni correction at an alpha level of 0.05, considering all genes tested across both variant filters (no filter and MAC ≤ 5): p ≤ 2.57e-06. FDR significance was similarly defined at the phenotype level, pooling p-values across both variant filters and applying a 5% false discovery rate using the Benjamini-Hochberg procedure. No gene reached FDR significance for MDD.

**Table S21. Single-variant association results for deleterious variants with MAC > 5 in genome-wide- and FDR-significant genes from the burden test analysis.** These results are extracted from the full single-variant analysis available in Tables S3–S7.

**Table S22. Minor allele frequency comparison for deleterious variants with MAC > 5 in genome-wide- and FDR-significant genes from the burden test analysis.** The table compares the MAF observed in Mision Origen with those reported in gnomAD, Regeneron Million Exome Variant Browser (RGC), All of Us (AoU), UK Biobank (UKB), and TopMed (Bravo). For each gene, the Mision Origen MAF corresponds to the variant with the strongest burden association result. When a variant was not present in an external data resource, MAF is reported as N/A. AF = allele frequency, AC = allele count. Allele counts/frequency estimates in RGC reflect probabilistic ancestry assignment based on individual haplotypes, and thus may appear as non-integer values.

**Table S23. Results from local ancestry-informed associations using Tractor-Mix.** In the table, AFR = African, AMR = Admixed American, EUR = European. Allele count threshold = 50 for all variants, besides 17:68433486:A:G (AC threshold = 10).

T**able S24. Conditional local ancestry-informed associations using Tractor-Mix.** In this table, AFR = African, AMR = Admixed American, EUR = European. Only run for variants in Table S23 with an allele count greater than 50 for all three ancestries.

**Table S25. Number of participants with EHR information at the time of analysis, stratified by recruitment site and diagnosis.** (a) Number of participants included in item-level phenotypic analyses and (b) MDD chart review are shown for total participants and 7:904169:T:TC variant carriers.

**Table S27. NLP-extracted item-level phenotypes.** All phenotypes that were reliably extracted from clinical notes, with their corresponding NLP method of extraction and endorsement by diagnostic group.

**Table S28. Model selection for item-level phenotypic analyses in participants with SMI and MDD.** Model selection in participants with (a) SMI and (b) MDD. For features with rare outcomes (fewer than 15% of individuals endorsing the feature; proportion_endorsing < 0.15), a Firth logistic model is used. Remaining models were chosen based on likelihood ratio tests. Models with LR_pval < 0.05 were modeled using zero-inflated negative binomial (ZINB).

**Table S29. Results from univariate, phenotypic analyses in participants with SMI and participants with MDD.** Effect of carrier status (non-carrier = reference) on item-level phenotypes are shown in participants with (a) SMI and (b) MDD. Models were adjusted for age of recruitment, sex, recruitment site, diagnosis (SMI only), and additional model specific covariates.

**Table S30. Univariate sensitivity analyses in participants with SMI + BDI, BD, BDI, and SCZ, for Bonferroni significant item-level phenotypes in participants with SMI.** (a) Model selection information for sensitivity analyses. Models were chosen based on likelihood ratio tests. Models with LR_pval < 0.05 were modeled using zero-inflated negative binomial (ZINB), otherwise, a negative binomial regression (NB) was used. Rare outcomes (fewer than 15% of individuals endorsing the phenotype; proportion endorsing < 0.15) were modeled using Firth logistic regression. (b) Results from univariate, sensitivity analyses in patients with SMI (main results), SCZ + BDI, BD, BDI, and SCZ

**Table S31. Sensitivity analysis: univariate item-level phenotypic analyses in participants with SMI and MDD, adjusting for the first five genetic PCs**. Model decision can be found in Table S28.

**Table S32. Best estimate diagnoses from MDD chart review, stratified by 7:904169:T:TC carrier status.** *One of the three participants with a best estimate diagnosis of BD, received the diagnosis after their recruitment date.

**Table S33. Results from chi-squared analyses comparing feature endorsement between MDD carriers and non-carriers.** Data obtained from manual chart review of 52 carriers and 52 non-carriers.
